# Optimizing TB policies using the global TB portfolio model: an economic analysis

**DOI:** 10.1101/2025.06.03.25328801

**Authors:** Srinath Satyanarayana, Sandip Mandal, Finn McQuaid, Sreenivas Nair, Suvanand Sahu, Nicolas A. Menzies, Sedona Sweeney, Rachel Sanders, Ines Garcia Baena, Richard G. White, Taghreed Adam, Mikaela Smit, Carel Pretorius

## Abstract

**Rationale:** Tuberculosis (TB) remains a global health crisis, disproportionately affecting low- and middle-income countries. Strategic resource allocation is essential to achieving the WHO End TB targets. Existing TB costing tools have limitations in conducting global analyses, prompting the development of a novel model tailored to address these gaps.

**Methods:** We developed a new, open-source TB costing model that simulates detailed TB care cascades comprising steps of screening, diagnosis, treatment, and prevention for those eligible - according to WHO guidelines for 20 distinct population groups. These include 10 groups each from patient-initiated and provider-initiated pathways, capturing variations in pulmonary status, age, HIV/ART status, and drug sensitivity. The model captures the cost of a large-scale vaccine. We demonstrate the model’s functionality through a case study that informed the Global Fund’s Investment Case for its 8th replenishment (2027–2029).

**Results:** In the case study, the model was first used to estimate the cost of implementing the TB Global Plan 2023 - 2030. This scenario incorporated intervention targets, normative standards of care, and the availability of new TB tools. An optimization routine applied to 29 high-burden countries estimated maximal TB impact under constrained funding scenarios. The results were also used to assess the potential impact and contribution of innovation within the Global Fund’s 8th replenishment.

**Discussion:** This new TB costing model offers improved representation of TB care complexity across diverse populations, with enhanced transparency, flexibility, and policy relevance. Its application in global TB strategy analysis highlights its value in informing investment cases and prioritizing interventions for maximal impact under resource constraints.

## Introduction

Tuberculosis (TB) remains one of the world’s deadliest infectious diseases, defying decades of control efforts and billions in global investment. Between 2002 and June 2024, the Global Fund (GF), the leading international funder of TB services in high burden settings, invested US$9.9 billion in TB programs to prevent and treat TB [1]. Annually more than US$4 billion is spent on TB programs when adding domestic spending and spending by other external donors to GF expenditures, with US$4.5 billion spent in 2023, as reported to the World Health Organization (WHO) [2], excluding health system costs. This is far below the US$22 billion needed annually for the 2023-2027 period to implement the TB Global Plan 2023-2030 [3] (TB GP), an amount committed to as part of the second political declaration to meet End TB targets 43] (by reducing TB incidence by 80% and TB deaths by 90% by 2030 relative to 2015 levels) following a United Nations High-Level Meeting (UNHLM) on TB in September 2023 [5]). This amount includes health system resource needs which represent about 11% of total TB GP resource needs. Recent reductions in U.S. government funding for global TB programs have further exacerbated this financial shortfall, highlighting the critical need for effective prioritization approaches [6], [7], [8], [9].

In this high-stakes environment, decision-makers must mobilize and allocate scarce resources with a clear understanding about the service levels that can be achieved with additional funding, and TB costing models are essential to guide these choices. When aligned with impact models, these tools enable policymakers to simulate different scenarios, compare strategies, and understand both the costs and health outcomes of interventions and packages of interventions, such as intensified case detection, improved treatment adherence or the large-scale rollout of new vaccines. Such models help bridge the gap between ambition and implementation by showing what it takes to reduce TB incidence and mortality.

A cornerstone of national strategic health planning efforts has been the OneHealth Tool (OHT) [10], a widely used, bottom-up TB costing model. It estimates the cost of TB programs by capturing both direct (e.g., diagnostics, treatment) and indirect costs (e.g., managing the TB program, staff training and conducting surveys). While OHT has proven valuable in country-level planning and even supported earlier efforts to cost the TB Global Plan, it presents significant limitations when it comes to costing global strategies, which is not its primary purpose. Its care cascade is not directly linked to specific population groups, making it difficult to tailor cost estimates to specific population and target groups, and its linkage to the TIME Impact model [11] does not allow for direct reflection of WHO guidelines or to efficiently support resource optimization. These shortcomings, while manageable in national applications, become significant obstacles in global strategy development.

These and other limitations of the OHT framework in global-level modelling applications, prompted calls from Global Fund and Stop TB technical guidance groups to improve and redesign key aspects of the model to inform the Global Fund’s Investment Case for the 8th replenishment 2027-2029 [12], as a first in a planned series of key applications of the new costing model. This analysis, which was published in 2025, is based on epidemiological and costing models for each disease (HIV, TB and malaria) and requires estimates for financial resource needs, optimal health impacts, return-on-investment (which is not directly modelled by this costing tool), and other consequences of using the resources that are to be mobilized.

OHT’s framework was improved, and a successor tool called the Integrated Health Tool (IHT) for planning and costing was developed [13]. This web-based tool provides planners with standardized methods to assess resource needs and costs, estimate health impact, do scenario comparisons and identify financing sources to meet costs. It follows a patient-centred approach to structure TB service delivery, allowing close alignment between the tool and WHO guidelines. It includes default estimates and cost inputs that can be adapted by the user. The costing component of the global TB portfolio model (GTPM) [14], the subject of this study, was developed in tandem with the Target Population (TP) component of the IHT tool (the part of the costing tool which manages target populations to be used in costing calculations), and its TP structure follows that of IHT closely. These and other design elements are discussed below.

This paper presents the methods and innovations behind this new global TB costing model, shares illustrative findings from its application, and explores the implications for resource needs estimation and future TB control strategies.

## Methodology

The Global Model Advisory Group was convened by the TB Modelling and Analysis Consortium (TB MAC), bringing together representatives from a range of organizations: including modelling experts affiliated with TB MAC, WHO, the Stop TB Partnership (STB), the Global Plan’s working group, the Global Fund’s guidance group, and others. The group met to discuss the limitations of existing costing tools for global-level TB analysis and to assess proposed improvements.

A new costing model was developed under the guidance of the advisory group to address these limitations and to meet emerging demands from the global modelling community (including those issued by STB’s and Global Fund’s guidance groups). It uses a bottom-up, ingredients-based approach to estimate the cost of different TB strategies

It is structured around screening algorithms and diagnostic methods that are aligned with WHO’s recommendation for appropriate screening, diagnosis, treatment and prevention. Approximately 70 interventions and services are organized into these algorithms, which are linked to specific target populations. Algorithms vary, in one or more aspects of the algorithms and by at least one variable among: age, pulmonary status, HIV status, drug-resistant TB (DR-TB) status, patient or provider-initiated service delivery, amongst other variables.

A new Target Population (TP) component was added to the costing component of GTPM. It computes both costing and impact inputs and serves to align the costing and impact models for a given scenario being modelled.

This mechanism as well as the calculation flow of the costing and impact models enabled the development of a simple prioritization method, which indicates service configurations that would optimize impact over the next GF replenishment period (2027-2029) under different allocation assumptions.

### Resource needs formula

For each population-algorithm pair, resource needs were determined by summing the algorithm ingredients and the underlying services, specifying the factors for each element:

**Resource needs(t) = Target population(t) x Population in need(t) x Coverage(t) x Unit cost(t) x Q(t),**

where:

● All the variables are time-dependent, with time (t) running between the base year and final year of a model projection.
● **Target population:** Interventions are linked to a population meant to receive the intervention or health service. For example, the target population for diagnosis with rapid molecular tests are all people eligible for testing (e.g. symptomatics, those of x-ray abnormalities and others).
● **Population in need (PIN):** PIN is a specification for a proportion of the population that is eligible for the intervention. For example, a proportion of all notified cases may need patient support.

Note that all interventions/services require the specification of a PIN value. Many PIN values are not directly based on data but are normative as per TB GP specification, for example that all diagnosis must be done with rapid molecular methods.

● **Coverage:** Service coverage targets are specified between 2023 and 2030. For example, diagnosis with rapid molecular tests may increase from 40% in the first year of a TB strategy to 100% by the final year of the strategy.
● **Unit costs as determined by ingredients:** This will typically comprise the following:

`o` Tradable components: commodities (e.g., all the commodities needed for a given diagnostic test)
`o` Non-tradable components: capital costs and staff time provided by technicians, doctors, community health workers and other types of staff.
`o` Costs resulting from outpatient and inpatient days are handled as part of the algorithms and included as separate unit cost components.
● **Quantity Q(t):** A quantity setting is used to control how many units of cost to apply for a given intervention. For example, multiple tests may be done during treatment.
● Coverage, PIN and intervention quantity settings were made consistent with Global Plan strategy targets.

### Direct and Indirect costs

Unit cost data came directly from the Value TB project [15] that have recently been made available for five countries, namely, Ethiopia, Georgia, India, Kenya and Philippines. These five countries serve as reference countries to which all other countries are linked, in a one-to-one mapping: Georgia was used as a reference for the upper-middle-income and high-income countries. India was used as a reference for the lower-middle-income/low-income countries in South Asia. The Philippines was used as a reference for the countries in the Western Pacific region. Kenya was used as a reference for the middle-income countries in Africa. Finally, Ethiopia was used as a reference for lower-income high-burden countries in Africa. Income-status classification is based on 2022 World Bank data.

For reference countries, local currency data for the country-specific base year and an inflate/convert methodology [16] were used to arrive at the unit costs for the base year2022. To extrapolate the unit costs from the reference country to the target country, an ingredients-based approach was used, as suggested by Torres-Rueda et al [17]. Each cost input in the ingredients-costing was classified as a tradable good (consumables), non-tradable good (overheads + capital costs) or staff cost. To convert the tradable goods from the reference country (R) to the target country (T), the cost of the tradable goods was converted into US$ in the base year. Tradable goods were inflated using US$-based inflation rates or taking the latest price from the Global Drug Facility (GDF) Medicines or Diagnostics Catalog [18], [19]. For extrapolating costs of non-tradable goods (NT) from a base country to the target country, the ratio of purchasing power parity [20] was used to obtain the equivalent costs in local currency for the base year, inflated in local currency using local country-specific inflation rates for the target year, and then converted back to US$ using the target year currency conversion rates. To convert staff costs (S) for a particular service from a base country to the second country, the staff time (in minutes) to conduct the activity and the estimated staff cost per minute in the base country were used. Staff time (in minutes) was extrapolated to the target country without any modifications from the base country. For converting the staff cost (per minute), the conversion rates from Serje et al. were used [21]. Gross Domestic Product (GDP) per capita multipliers and the nominal GDP ratios were used to convert the staff wages per minute from the base country base year to the target country base year; this cost was multiplied by country-specific inflation rates, and the target year staff cost values in the local currency were obtained and then converted to US$ in the target year conversion rates. The total unit cost in the target country for the target year is the sum of T, NT and S.

For the cost of medicines (for the treatment and prevention of TB disease) and consumables, the prices in US$ from the latest GDF Diagnostics and Medicines Catalogs were used. It was estimated that Computer Aided Detection (CAD) would cost an additional US$ 1 per person (assuming high volumes) undergoing digital chest X-ray.

For all unit cost calculations, the latest country-specific GDP deflation and US$ conversion rates published by the World Bank [22], [23] were used to adjust the inflation and currency conversions from the base year (2022) to the target year of the TB GP (2030).

After costs were converted to 2022 US dollars, a US GDP deflator was used to project unit costs from 2023 onward.

Expenditure data reported to WHO are used to estimate indirect costs as a markup of the cost of direct services (diagnosis, treatment and so on) represented by “programme costs”, or costs above the patient level. Averaged over the countries that report TB expenses to WHO, this cost is more than 50% of direct costs.

In TB Global Plan applications an additional markup was added to overcome a lack of investment in key enabling activities. Following the recommendation of the Global Plan task force a uniformly fixed-percentage markup was added for specific “enabling” activities, including direct patient support (5%), advocacy & communications (1%), Community, Rights & Gender (CRG) (6%, based on NSP budgets of programmes implementing CRG) and Public Private Mix-TB (PPM-TB) (12%, for countries with a high proportion of people seeking care with private-sector care providers).

Detailed budgets of a few countries, such as Democratic Republic of Congo, Georgia, India, Philippines and Tajikistan, judged to be representative in terms of budgeting for enabling activities were used to estimate the size of enabling cost categories.

The list of interventions included in the costing model, and their respective unit sources, is shown in Table 1 of the Appendix. Services fall into 11 categories: testing for drug-sensitive (DS) and drug-resistant (DR) TB, testing for TB infection, tests during treatment monitoring and follow-up, other tests and procedures, non-regimen treatment-related costs, regimens for treating active paediatric TB, regimens for treating active adult TB, post treatment services, regimens for TB prevention, vaccination costs and other costs.

Key elements of the care-cascade are calculated in the TP component, detailed below. These elements include the number screened or clinically assessed for TB disease, the number referred for TB diagnosis, the number initiated on TB treatment, the number diagnosed with active TB, the number eligible for TB preventive treatment (TPT) and the number initiated on TPT.

The model estimates the target population for a large-scale roll out of a new vaccine among the population 10 years and older. The model achieves and then maintains the stated annual coverage of the vaccine in the population 10 years and older. Once the stated final scale-up level is achieved it is maintained: a) by vaccinating enough individuals to account for the balance between those entering an age category (by aging into it) and those leaving it (by aging out of it or by death) and b) vaccinating individuals entering the 10-year-old age group.

### The Target Population component

A target population (TP) component of the costing model was developed to facilitate calculation of the care cascade and to interface between the costing and the impact model, thereby ensuring good alignment between impact and cost scenarios.

To do this, the TP component is designed to estimate the various target populations needed to cost the algorithms for approximately 20 patient population-based algorithms. Essentially it processes the care cascade for each patient population by using inputs that define population size, TB infection and disease prevalence, a screening algorithm (both the algorithm and its coverage) and a diagnostic method, with an estimated sensitivity and specificity for each. TB prevention is also explicitly handled through inputs for TB eligibility (presumptive or following a test for TB infection using tuberculin skin test (TST) or interferon-gamma release assay (IGRA) according to recommendations that are tailored to patient groups). These elements are described in more detail in the section: *General structure of the Target Population model*, in the Appendix.

To capture the necessary variations in screening and diagnosis, populations at risk of TB disease are differentiated according to how they access services, namely patient-initiated (10 populations) or provider-initiated (10 populations) screening, and by properties that influence intervention details such as age, pulmonary and HIV status. In total there are 20 population or patient groups at risk of TB disease, as detailed in Table 2 of the Appendix.

Diagnosed and notified patients are split according to pulmonary status, age and HIV status using data obtained from the WHO’s case notifications database. This so-called notification split is based on the final year of WHO data, and it is applied to the user-input notification target to obtain projections beyond the final year of WHO data for different patient types.

Screening volumes for provider-initiated populations are directly calculated, and it is back-calculated for patient-initiated populations. Meaning, the method starts with the number of each patient type, that remains after removing patients from provider-initiated programs from the total, and then, based on the selected clinical evaluation algorithm and an assumed prevalence of TB in a patient population, calculate how many persons must have been screened to result in the final notified number. This number cannot be obtained directly from a notification split explained in the previous section because the notifications database from WHO does not indicate the population groups linked to notifications – it reports the total number of notifications only.

The total number of patients diagnosed and initiated on treatment for TB disease is determined by aggregating patient types over all the target population variations by age and pulmonary status. The group that a notified patient comes from: patient initiated, household contacts, PLHIV on ART or high-risk groups, plays no role in the selection of appropriate regimen used for treatment. Treatment details vary according to age, resistance profile and severity of disease, as detailed in the Appendix. The flow of information through patient-specific care-cascades in the Target Population component of GTPM is shown in Figure 1. Patient groups that are handled in both patient- and provider-initiated groups are shown in green. Their overlap is explicitly handled in the calculation of the number of patients screened and diagnosed in patient-initiated groups.

**Figure 1:**
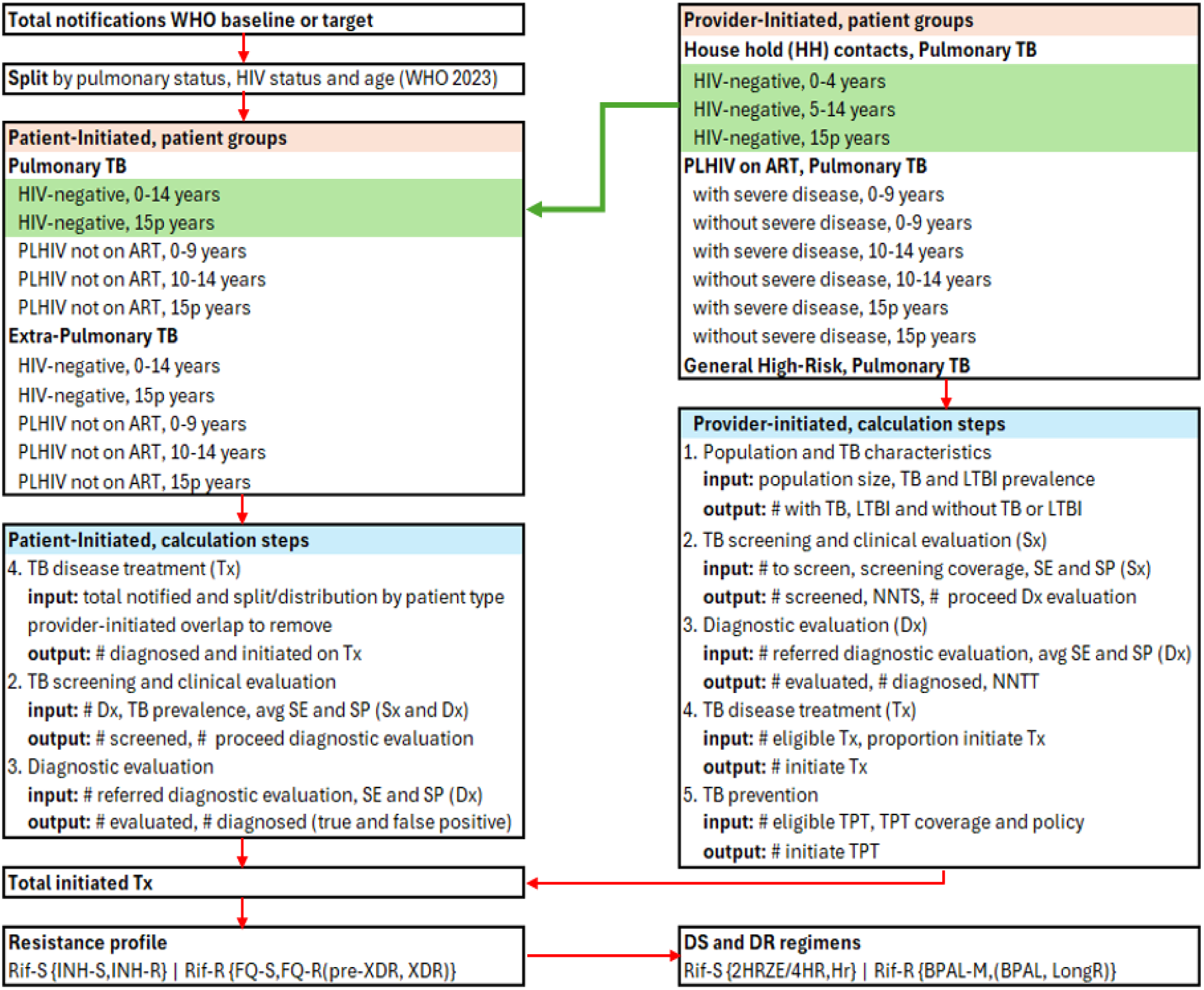
Schematic illustration of the flow of information of the costing component of GTPM, from a total notification target to volumes for screening, diagnosis, treatment and prevention. Abbreviations related to calculation flow: # - number, Sx – screening, Dx – diagnosis, LTBI – TB infection, SE – sensitivity, SP – specificity, Tx - treatment. The green line indicates overlap between patient- and provider-initiated programs with respect to total notifications. Abbreviations related to a resistance profile (simplified): Rif-S-Rifampicin sensitive, INH-S-Isoniazid sensitive, INH-R – Isoniazid resistant, Rif-R – Rifampicin resistant, FQ-R Fluoroquinolone sensitive, FQ-Fluoroquinolone resistant, pre-XDR, XDR-pre-XDR and resistant to at least one of Bedaquiline, Linezolid, Levofloacin, Moxifloxacin (see Figure 2, Appendix). Abbreviations related to regimens linked to resistance profile: 2HRZE/4HR, Hr, BPAL-M, BPAL, Long-term regimens (see Table 1, Appendix)

Figure 2 shows a further structuring of the model. Services are grouped into (blue column) those related to screening, diagnosis and other tests, (green column) treatment including regimens, monitoring and other services provided during treatment, and (brown column) prevention including regiments for adults and children, monitoring and other services provided during preventive treatment. These inputs are listed in Appendix Tables 3-16, according to the Table mapping given in Figure 2. Regimen costs were adapted from GDF, meaning they were mapped to a single average unit cost per regimen, using parameters such as age and treatment duration.

**Figure 2:**
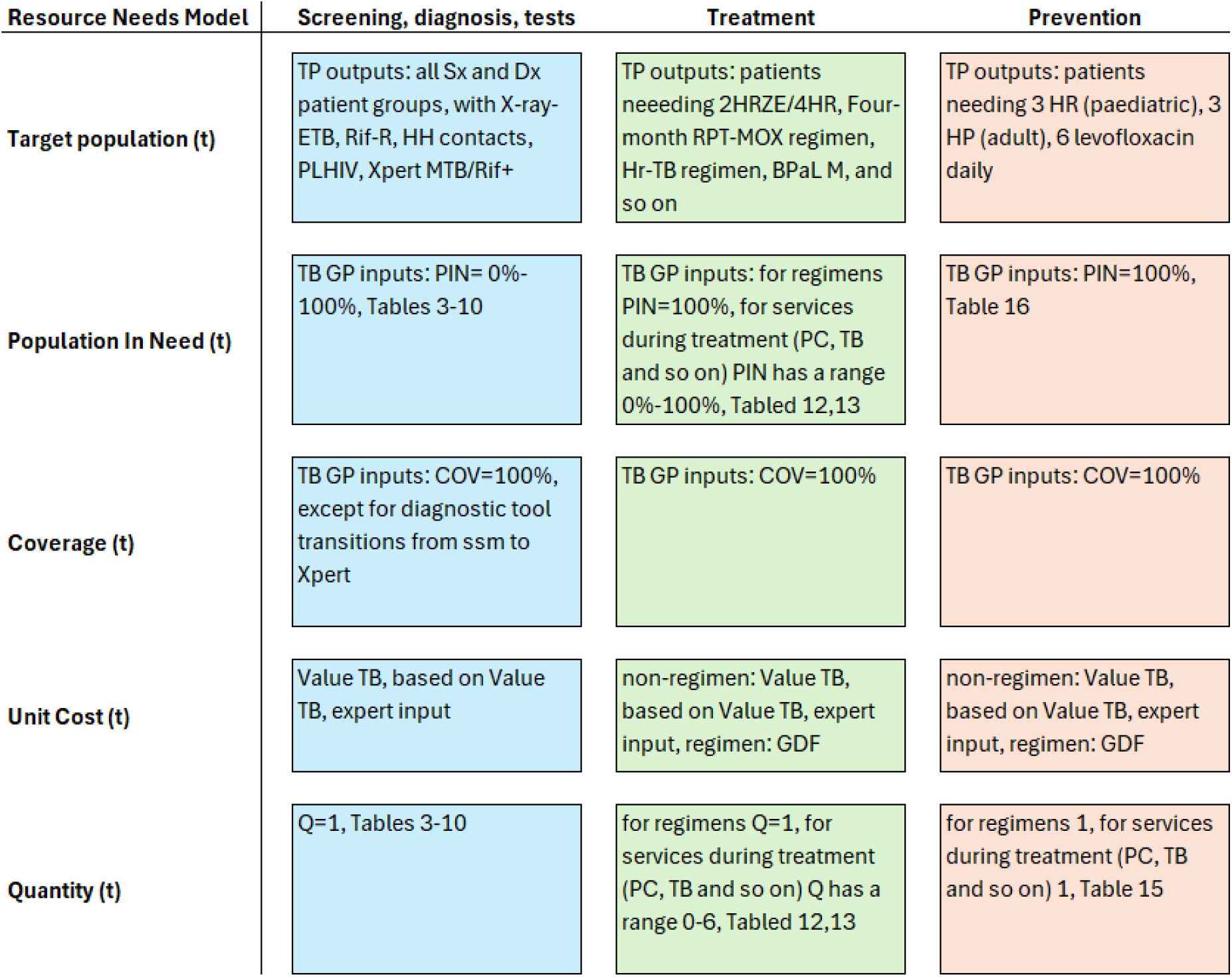
Schematic illustration of the costing model structure, showing at high-level how Population in Need(t), Coverage(t), Unit Cost(t), and Quantity(t), each of these a function of time t, is arranged by service area: screening, diagnosis and all tests (blue), treatment for TB disease (green) and TB prevention (brown). Abbreviations: TP-Target Population component, TB GP-TB Global Plan 2023-2030, GDF – Global Drug Facility. Table numbers refer to tables in the Appendix.

### Impact from the Target Population Component

In addition to estimating target population sizes, the Target Population (TP) component projects service volumes for screening and diagnosis, TB prevention, and treatment under a given scenario. The ratio of service volumes between two scenarios, typically comparing a scale-up to a baseline or constant coverage scenario, is then used to adjust specific baseline parameters within the transmission component of GTPM [14] related to service delivery and intervention effects.

For instance, a detection ratio between a baseline and scale-up scenario is calculated based on the algorithmic details of all patient populations modelled within the TP component. These computational details incorporate the patterns of screening scale-up, the sensitivity and specificity of the screening algorithms and diagnostic tools, and the estimated or assumed prevalence of TB disease within the population.

Impact ratios for TB prevention are similarly generated by comparing the number of individuals receiving tuberculosis preventive therapy (TPT) across baseline and scale-up scenarios, with greater weighting given to high-risk groups in the calculation.

Treatment outcome ratios are derived from comparisons of drug susceptibility testing (DST) coverage and default rates. These calculations also account for reductions in treatment success rates among second-line (SL) patients who receive inappropriate first-line (FL) regimens due to the absence of DST.

To estimate the impact of the scenario projection, hazard ratios for a given scenario compared to a counterfactual, quantify increased detection (**Hd** in Figure 1), prevention (**Hi** in Figure 1) and treatment outcomes (**Htsr** in Figure 1) are used to change impact related parameters in the transmission model [14]. The flow of information between the TP component and the transmission and costing components of GTPM is shown in Figure 3, which is taken from [14].

**Figure 3:**
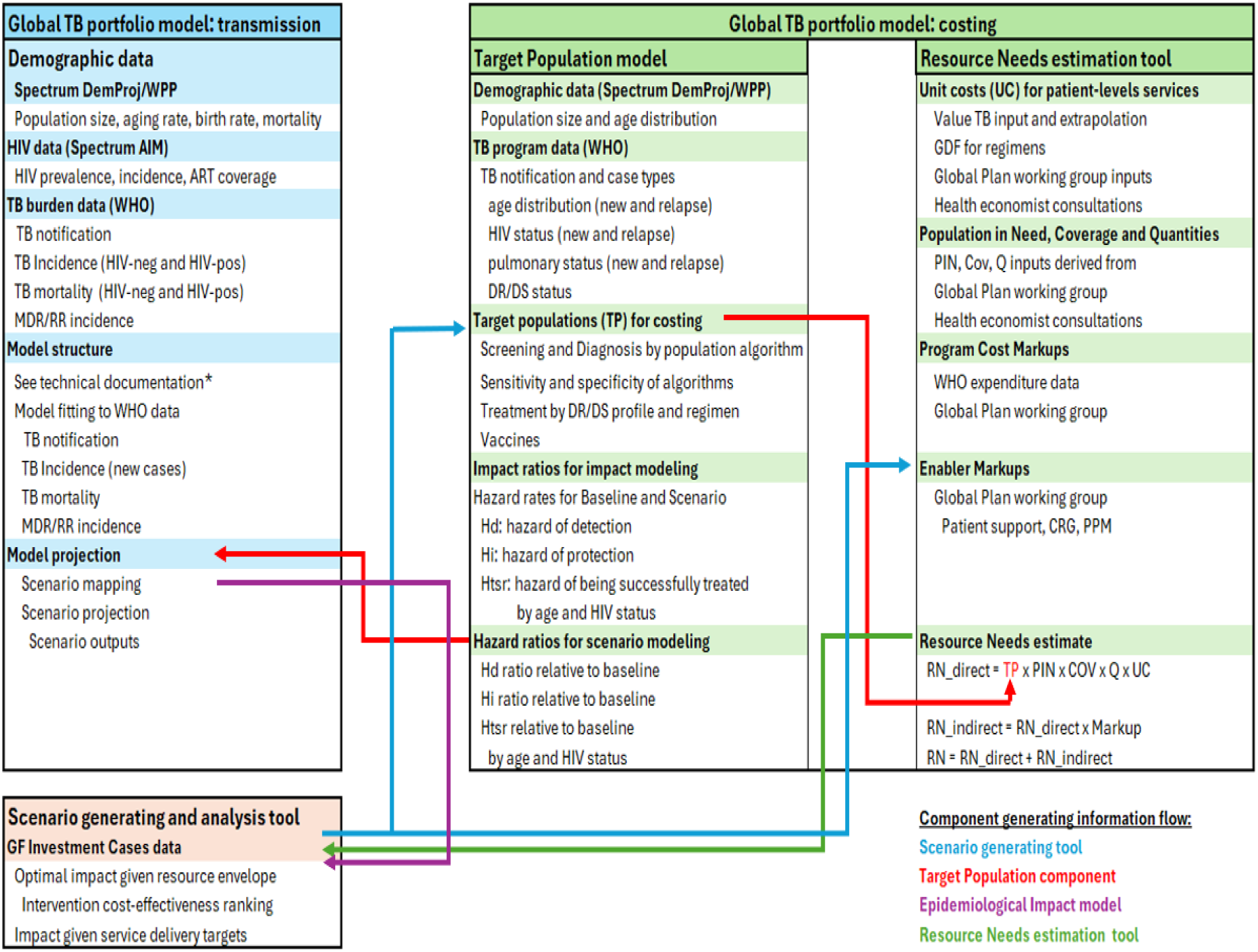
Schematic illustration of the flow of information between the Target Population model, the transmission and costing component of GTPM. RN-resource needs, PIN-population in need, COV-coverage, Q-quantity, UC-unit cost. *Details of the GTPM model’s transmission component can be found in [14].

In an application such as Global Fund’s Investment Case analysis, a scenario-generating tool is used to populate the TP structure with intervention details (comprising baseline and target coverage levels) corresponding to a specific scenario. Hazard ratios are applied to a calibrated epidemiological impact model and TP outputs (volumes or patient numbers requiring services needed for costing) are used to calculate resource needs. Results from these models are used to produce optimal impacts of resource allocations, return on investment results and other outputs needed for a given analysis.

### Resource prioritization

The Global Plan to End TB 2023–2030 provides an ambitious intervention framework aimed at ending TB by 2030 in an unrestricted resource environment. However, the resources required for its full implementation are substantial and cannot be achieved without ongoing, large-scale resource mobilization efforts. In most countries, it is anticipated that domestic and external financing will not fully meet the resource needs for implementing the TB GP. Recent reductions in U.S. government funding for global TB programs have further exacerbated this financial shortfall, highlighting the critical need for effective prioritization approaches.

For previous Global Fund Investment Case analysis our modelling tools could not support optimization and prioritization for global analyses due to runtime constraints and the absence of a prioritization framework that is both informative and capable of accounting for dependencies among interventions. This functionality has now been enabled through a redesign of the costing and impact models, their shared interface with the TP model, and the development of an ancillary scenario generation and analysis tool. This new scenario generation tool constructs intervention scenarios and coordinates the interaction between the TP, cost, and impact models.

The prioritization algorithm begins by establishing a baseline scenario that reflects a funding-constrained environment. Program elements (interventions) are then sequentially added in order of increasing cost-effectiveness until the available budget is fully allocated. These interventions are modelled at the level of the Global Fund’s TB “modules,” with each module encompassing multiple activities [24]:

● Increase detection by scaling up systematic screening. Note this implies that baseline notification is predominantly driven by passive case finding.
● Scaling up DST systematically towards universal DST. This will also improve treatment success.
● Scale up TB prevention (TPT) within provider-initiated or systematic screening programs, namely within HH contact tracing, PLHIV on ART and general high-risk groups.

### Constructing a cost-impact curve

The next step of the algorithm is to estimate a cost-effectiveness measure or objective function for each of these broad intervention packages. This is done by scaling each package from baseline to TB GP levels in turn while keeping the other packages at baseline levels. Three intervention packages are included: Expanding systematic screening (labelled ACF in Figure 1), Expanding DST (labelled DST&LTFU in Figure 1) and Expanding Prevention (labelled TPT in Figure 1) with the addition of the last package reaches the full TB GP 2023-2030. The total number of new TB cases and TB deaths, the objective function to minimize) is estimated over a given period, namely 2027-2029 for the next GF replenishment.

There is no simple way to estimate minimally detrimental outcomes when resources drop below baseline levels and when baseline notification levels are not reached. We take the simplifying approach of modelling resource needs at 0%, 25%, 50% and 75% of baseline notification levels, assuming the same relative distribution of patients (by age, by bacteriological confirmation status, by resistance profile, etc) as the baseline distribution of these variables.

Thus, three sections of a cost-effectiveness curve are produced: Below baseline resources, baseline resources and successive addition of systematic screening, DST and TPT to baseline service delivery, resources up to the requirement of the implementation of the full TB GP.

## Results

### Cost-effectiveness curves

A typical cost-effectiveness curve produced by GTPM is presented in Figure 4, showing a maximal impact that could result from a range of resource availability considerations. While not sufficiently informative to guide prioritization at country-level, it provides a measure of maximal impact which here refers to the lowest number of TB deaths and cases, given a TB resource cap. The method can also maximize averting a weighted combination of TB deaths or cases, or TB deaths or cases alone. In the example below, equal weight is given to TB deaths and cases.

**Figure 4:**
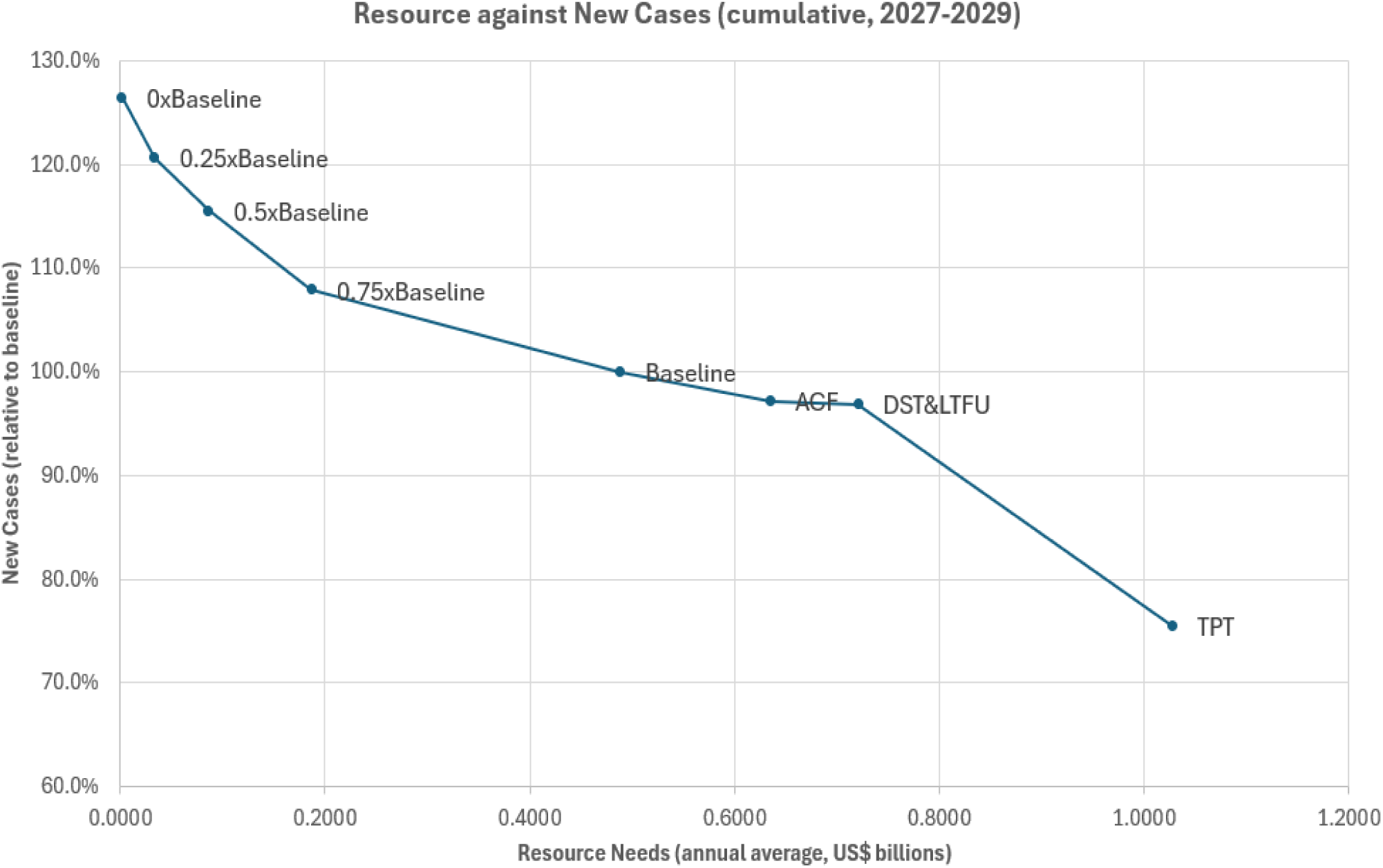
Optimal cost-effectiveness curve for Indonesia, the x-axis value for TPT corresponds to the implementation of the full TB GP 2023-2030.

Given the uncertainties with the underlying assumptions and resulting location of points on this curve, a recommendation is followed regarding sharing of results in global strategy reports or the Investment Case analysis, that the interventions comprising the curve should not be mentioned individually, as it could lead to confusion, such as defunding one of ACF, DST or TPT (or an intervention within each of these packages of interventions) in favour of expanding another. Global Fund’s considerations are wider than cost-effectiveness alone in their detailed guidance for TB prioritization at country level, which accounts also for equity and sustainability, amongst other considerations.

### Global Fund Investment Case analysis

#### Optimal Impact

Several key estimates were produced with the costing component of GTPM, informing the Investment Case of the 8th Global Fund replenishment [12]. Foremost is the estimate of the total resource requirement for 2027-2029 which is based on the TB Global Plan 2023-2030. There are important deviations from the Global Plan in the Investment Case scenario. First, service volumes are set from performance targets for the pre-replenishment period (2023-2026). Coverage is then scaled-up over the replenishment period (2027-2029) with agreed adjustments made to the Global Plan coverage levels to delay the proposed large-scale rollout of new vaccines to begin in 2029. The overall implication of these scale-up patterns is a delay to maximal service delivery coverage relative to the TB Global Plan scenario, which assumes a highly optimistic scale-up pattern and the availability of resources to enable it. This version of the Global Plan scenario, and its intervention packages, is used to construct the cost-effectiveness curve shown for Indonesia, in Figure 4. Maximal impacts are then obtained by using a proposed resource envelope and a maximal impact value for new TB cases and deaths is then interpolated on this curve. This establishes an optimal scenario and its projected impacts, relative to the baseline scenario (of continuing coverage from 2023 forward). In total 29 country models were developed for the analysis, out of the 30 highest burden TB countries in the GF’s TB portfolio. The Democratic People’s Republic of Korea, a country in the list of 30 high-burden countries, did not have sufficient data for model calibration. The 29 countries with calibrated models represent 90% of the TB burden among GF eligible countries.

The impact results show, in Figure 5, that the proposed replenishment would make significant progress towards reaching the TB Global and End TB impact targets but will reach them by 2035 instead of 2030, even if resources are used optimally by all countries in the GF’s TB portfolio, in the general sense assumed by the optimization method of the model, leading to maximal impact. The model further shows that by 2035 impact reached would be much greater relative to 2035 impact targets, as by then all interventions (based on current and new tools) are brought to scale, including the rollout of an effective but costly large-scale vaccine.

**Figure 5:**
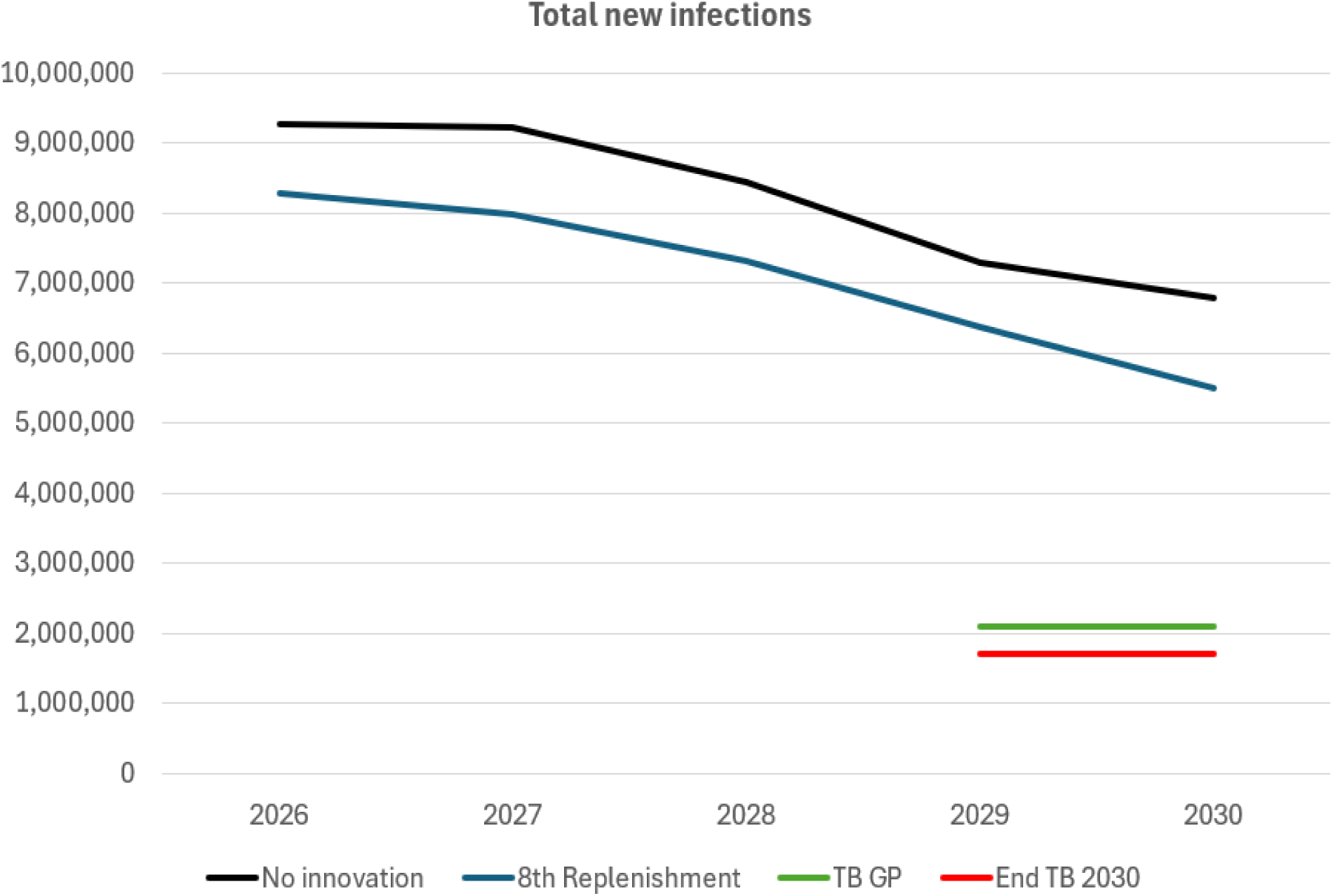
The maximal impact of the Investment Case is shown in the blue line. A counterfactual Investment Case scenario with no innovation from 2023 onward is shown in the black line. The level of new infections reached following the implementation of the Global Plan is shown in green and the End TB incidence target for 2030, in the red line.

#### Impact of Innovation on the Global Fund’s Investment Case

Innovation in TB programs is critical to achieving TB control targets as shown in the TB GP. Its role in the current (7^th^) and next (8^th^) replenishment cycles was studied in detail. The TB GP demonstrated the cost of delaying the implementation of the TB GP, and of delaying investments in R&D needed to develop and utilize new tools for diagnosis, prevention and treatment to end TB. To estimate the impact of recent innovation on the 8^th^ replenishment, we identified key interventions that are currently available, including pediatric diagnosis and treatment, improved TPT regimens, improved SL treatment, molecular diagnostic tools, and engaging care providers (PPM) as key innovative elements of TB programs. Tools that are expected to become available include a large-scale vaccine. These elements were removed from all country intervention structures from 2023 onward to approximate a counterfactual where no recent innovation was acting. The area between the black and blue line in Figure 5 represents the potential impact of innovation on the 8^th^ replenishment.

## Discussion

This study introduces a novel TB costing model developed to address critical limitations of existing frameworks, such as the OneHealth Tool, in costing global TB strategies. The model provides a flexible, high-fidelity platform for estimating resource needs for the implementation of national and global TB control strategies. Importantly, it captures key variations in TB service delivery relevant to normative costing exercises led by the Stop TB Partnership and the Global Fund, including differences across the TB care cascade for pulmonary and extrapulmonary TB, by age group, HIV and ART status, and across patient-versus provider-initiated pathways.

A major strength of this costing model lies in its open-source design, which promotes transparency, reproducibility, and adaptability to both global and country-specific implementation structures. This framework facilitates efficient updates to core assumptions—such as unit costs, populations in need, service quantities, and other key inputs—ensuring the model remains aligned with evolving technical guidance and data.

The prioritization algorithm developed alongside the costing tool supports prioritization at the global or portfolio level but is not designed for detailed country-level optimization. Although resource optimization was not required for the TB Global Plan analysis—given its ambitious and normative scope—the prioritization algorithm was employed to estimate the maximum potential impact of the Global Fund’s Eighth Replenishment, which aims to mobilize US$18 billion [12]. As part of this investment case analysis, the model also estimated the potential contribution of innovation—defined as a specified set of new and existing tools—to the overall impact achievable through replenishment resources.

Currently, the model does not support fine-grained country-level optimization decisions, such as whether a country should prioritize systematic screening and prevention or focus on enhancing passive case-finding among symptomatic individuals. However, the model and prioritization methods can be adapted for country-level application, provided sufficient expert input is available to customize the Global Plan intervention framework to national contexts. Such refinements are likely to become increasingly important given the tightening financial environment for TB programming, exacerbated by recent reductions in U.S. government support for global TB efforts.

Several limitations of the model must be acknowledged. As with most TB costing exercises, the model relies heavily on assumptions and expert judgments due to the limited availability of country-specific data. These include extrapolations from the Value TB study to countries not participating in the Value TB project, assumptions regarding the proportion of TB patients requiring specific services (e.g., psychosocial support, advanced diagnostics, or surgical interventions), and estimations of service delivery mark-ups for models such as public-private mix (PPM) and community engagement activities. Indirect costs are modelled as proportional to direct costs, despite uncertainties regarding how programmatic overheads scale with caseloads. Such limitations reflect broader data gaps in the field of TB costing and are not unique to this model.

There are several promising directions for further improving the costing tool. One key area is to incorporate the cost and impact of interventions targeting TB determinants—such as undernutrition, smoking, and diabetes—which are emphasized both in the End TB Strategy [3] and the TB Global Plan [5]. Accurately modelling these interventions will require technical guidance from global partners, including STB and WHO, particularly regarding the definition and scope of interventions. For example, defining an intervention for undernutrition would require specifications on caloric and nutritional content as well as guidance on the recommended duration of support.

In conclusion, the new TB costing model represents an important step forward in supporting strategic planning and resource allocation for global TB control. Further refinement, including the integration of TB determinant interventions and enhancements for country-level application, will strengthen its utility in increasingly resource-constrained environments.

## Conclusions

In conclusion, this costing component of the global TB portfolio model represents a significant step forward in producing model-based information towards TB resource needs estimation and resource mobilization. It provides a robust analytical framework to estimate resource needs of diverse intervention packages, it allows investment prioritization, and it guides policy and policy implementers toward meeting the WHO’s End TB targets.

## Supporting information

Technical Appendix

## Data Availability

All data produced in the present study are available upon reasonable request to the authors. Key data sources are shared in a GitHub respository

https://github.com/CarelPretorius/GlobalTBCostingModel

## Acknowledgements

The authors acknowledge the global portfolio model advisory group convened by the TB Modelling and Analysis Consortium (TBMAC), with representatives from various organizations, including costing experts linked to TBMAC, WHO, STB and the Global Plan’s working group, Global Fund’s guidance group and others whose inputs on the model design, functionality and validation have greatly helped to improve the costing component of the global portfolio TB model.

