## Supplementary material for "Optimizing TB policies using the global TB portfolio model: an economic analysis": Technical Appendix

This document details the technical description of the TB care algorithms used for estimating the resource needs of the TB Global Plan 2023-2030. The costing component of the global TB portfolio model can be configured to estimate the cost of other algorithms, as needed.

#### **Data sharing**

Key databases and code for key functions can be found at:

https://github.com/CarelPretorius/GlobalTBCostingModel

#### **List of interventions/services and source of unit cost**

The list of interventions included in the costing model is shown in Table 1. Each intervention is shown with its “Method”, which indicates 1) “Value TB”: if it is based on the Value TB extrapolation method directly; 2) “From Value TB”: if its consumables are given as direct input (in the TB GP it is provided by the GP working group), but its non-tradable goods are based on a comparable Value TB unit cost; and 3) Input (Expert): if it is specified as a single value or “Lump sum”, such as all the treatment costs from the GDF Catalogs.

*Table 1: List of services, their explanation and the source of Unit cost*

| **Service/intervention code** | **Explanation** | **Method/Source** |
| --- | --- | --- |
| **Testing for DS- and DR-TB** |  |  |
| OPD-Scr | screening visit to health facility | Value TB |
| OPD-Dx | diagnostic visit to health facility | Value TB |
| SCT | sputum/specimen collection and transportation | Value TB |
| CXR | chest X-ray | Value TB |
| CAD | computer-aided detection | Input (Expert) |
| P-CXR | portable-chest X-ray | Input (Expert) |
| GA | gastric aspiration (for collecting specimens for diagnosis of TB in children <5 years of age) | Value TB |
| LF-LAM | lateral flow urine lipoarabinomannan assay | Value TB |
| mWRD-1 | molecular WHO-recommended rapid diagnostic test ( Truenat® MTB test) | Value TB |
| mWRD RR-1 | molecular WHO-recommended rapid diagnostic test for detection of rifampicin resistance (Xpert MTB/Rif Test) | Value TB |
| mWRD-2 | molecular WHO-recommended rapid diagnostic test for detection of rifampicin resistance (Truenat® MTB Rif test) | Value TB |
| mWRD-3 | molecular WHO-recommended rapid diagnostic test for detection of resistance to first- and second-line drugs (Xpert MTB/XDR) | Value TB |
| LC | liquid culture | Value TB |
| CV | community visit | Value TB |
| LPA-FLD | line probe assay for first-line drug | Value TB |
| LPA-SLD | line probe assay for second-line drug | Value TB |
| CRP | c-reactive protein | Value TB |
| TGS | targeted gene sequencing | Input |
| FNAC | fine needle aspiration cytology | Value TB |
| CT-scan | computed tomography scan | Value TB |
| **Testing for LTBI** |  |  |
| CT visit | contact tracing visit | Value TB |
| TST | tuberculin skin test | Value TB |
| IGRA | interferon-gamma release assay | Value TB |
| TBST | TB antigen-based skin tests (TBSTs) | Value TB |
| **Treatment monitoring and follow up** |  |  |
| TM | treatment monitoring | Value TB |
| OPD treatment | outpatient department treatment visit | Value TB |
| SSM | sputum smear microscopy | Value TB |
| SSC | sputum culture monthly | Value TB |
| FU-PTT | follow-up post-TB treatment | Value TB |
| FU-TT | follow-up during TB treatment | Value TB |
| LTFU tracing | tracing those who are lost to follow-up | Value TB |
| **Other tests and procedures** |  |  |
| HIV-Dx | HIV diagnostic testing | Value TB |
| DM | diabetes mellitus | Value TB |
| RFT | renal function test | Value TB |
| ECG | electrocardiogram | From Value TB |
| LFT | liver function test | Value TB |
| SGPT | serum glutamic pyruvic transaminase | Value TB |
| SGOT | serum glutamic-oxaloacetic transaminase | Value TB |
| Biopsy | biopsy | From Value TB |
| USG | ultrasonography | Value TB |
| MRI | magnetic resonance imaging | Value TB |
| **Treatment-related** |  |  |
| DOT | directly observed treatment | Value TB |
| DAT | digital adherence technology | Input (Expert) |
| PSC | patient support costs | Input (Expert) |
| PC | patient counselling | Value TB |
| BeddayDS | In patient management of drug-sensitive TB | Value TB |
| BeddayMDR | In patient management of drug-resistant TB | Value TB |
| aDSM | active TB drug safety monitoring and management | Value TB |
| AE | adverse event management through IP care | Value TB |
| Surgery | elective partial lung resection (lobectomy or wedge resection) | Input (Expert) |
| **Children/paediatric: regimens for treating active TB** | |  |
| 2HRZE/4HR (pediatric) | Treatment of Pulmonary TB pediatric six-month TB treatment regimen containing isoniazid, rifampin, pyrazinamide and ethambutol for two months/isoniazid plus rifampin for four months | Input (GDF) |
| 2HRZ/4HR (pediatric) | 6-month TB regimen for children | Input (GDF) |
| 6HRZEto (Pediatric) | In children and adolescents with bacteriologically confirmed or clinically diagnosed TB meningitis (without suspicion or evidence of MDR/RR-TB), a 6-month intensive regimen (ETO= Ethionamide) | Input (GDF) |
| 4-month shorter regimen [2HRZ(E)/2HR] | In children and adolescents between 3 months and 16 years of age with non-severe TB (without suspicion or evidence of MDR/RR-TB), a 4-month treatment regimen (2HRZ(E)/2HR) should be used. | Input (GDF) |
| Hr-TB regimen (pediatric) | pediatric six-month regimen for rifampicin-susceptible and isoniazid-resistant TB. rifampicin, ethambutol, pyrazinamide and levofloxacin | Input (GDF) |
| Short all-oral BDQ regimen (pediatric) | shorter all-oral bedaquiline-containing regimen for MDR-/RR-TB of 9–12 months’ duration | Input (GDF) |
| BPAL-M (pediatric) | bedaquiline, pretomanid and linezolid + Moxifloxacin regimen (6-9 months) | Input (GDF) |
| Delamanid-based regimen (pediatric) | pediatric treatment regimen for MDR-TB or XDR-TB containing delamanid | Input (GDF) |
| Longer DR-TB regimens (pediatric) | TB treatment regimen for DR TB, which may last 18-24 months | Input (GDF) |
| **Adults: regimens for treating active TB** | |  |
| 2HRZE/4HR | six-month TB treatment regimen containing isoniazid, rifampin, pyrazinamide and ethambutol for two months/isoniazid plus rifampin for four months | Input (GDF) |
| Four-month RPT-MOX regimen | four-month rifapentine-moxifloxacin regimen for the treatment of DS pulmonary TB | Input (GDF) |
| Hr-TB regimen | six-month regimen for rifampicin-susceptible and isoniazid-resistant TB | Input (GDF) |
| BPaL M | regimen of bedaquiline, pretomanid and linezolid + Moxifloxacin for 6–9 months | Input (GDF) |
| BPaL | regimen of bedaquiline, pretomanid and linezolid for 6–9 months | Input (GDF) |
| Short all-oral BDQ regimen | shorter all-oral bedaquiline-containing regimen for MDR-/RR-TB of 9–12 months’ duration | Input (GDF) |
| Longer DR-TB regimens | TB treatment regimen for DR TB, which may last 18-24 months | Input (GDF) |
| Long regimen for DR-TB, containing delamanid | TB treatment regimen for MDR-/RR-TB containing delamanid, which lasts at least 18 months | Input (GDF) |
| **Adults and children: Post TB treatment** | |  |
| PTLD care | post TB lung disease care | Input (Expert) |
| Palliative care | palliative care | Input (Expert) |
| **TB Preventive treatment** |  |  |
| 3 HR (pediatric) | prevention, children | Input (GDF) |
| 3 HP (adult) | prevention, adults | Input (GDF) |
| 6 levofloxacin daily (pediatric) | preventive treatment of MDR-TB, children | Input (GDF) |
| 6 levofloxacin daily (adults) | preventive treatment of MDR-TB, adults | Input (GDF) |
| **Vaccine** |  |  |
| Vaccine | TB vaccine | Input (Expert) |

##### **Target Population patient groups**

The population groups were classified as either patient initiated (passive case finding) or provider initiated (active/systematic screening).

*Table 2: List of population groups*

|  | **Group Description** |
| --- | --- |
| **Patient Initiated groups** |  |
| 1 | Pulmonary TB: HIV-negative, Children aged < 15 years |
| 2 | Pulmonary TB: HIV-negative, Adults 15 years and above |
| 3 | Pulmonary TB: PLHIV not on ART, Children aged 0 to 9 years |
| 4 | Pulmonary TB: PLHIV not on ART, Children aged 10 to 14 years |
| 5 | Pulmonary TB: PLHIV not on ART, Adults aged 15 years and above |
| 6 | Extra-Pulmonary TB: HIV-negative, Children aged < 15 years |
| 7 | Extra-Pulmonary TB: HIV-negative, Adults 15 years and above |
| 8 | Extra-Pulmonary TB: PLHIV not on ART, Children aged 0 to 9 years |
| 9 | Extra-Pulmonary TB: PLHIV on ART, Children aged 10 to 14 years |
| 10 | Extra-Pulmonary TB: PLHIV not on ART, Adults aged 15 years and above |
| **Provider Initiated groups, Household (HH) contacts** |  |
| 11 | HH Contacts, Pulmonary TB: HIV-negative, Children aged 0 to 4 years |
| 12 | HH Contacts, Pulmonary TB: HIV-negative, Children aged 5 to 14 years |
| 13 | HH Contacts, Pulmonary TB: HIV-negative, Adults aged 15 years and above |
| **Provider Initiated groups, PLHIV on ART** |  |
| 14 | CLHIV, Pulmonary TB: PLHIV on ART with severe disease, Children aged 0 to 9 years |
| 15 | CLHIV, Pulmonary TB: PLHIV on ART without severe disease, Children aged 0 to 9 years |
| 16 | CLHIV, Pulmonary TB: PLHIV on ART with severe disease, Children aged 10 to 14 years |
| 17 | CLHIV, Pulmonary TB: PLHIV on ART without severe disease, Children aged 10 to 14 years |
| 18 | PLHIV, Pulmonary TB: PLHIV on ART with severe disease, Adults aged 15 years and above |
| 19 | PLHIV, Pulmonary TB: PLHIV on ART without severe disease, Adults aged 15 years and above |
| **Provider initiated high-risk clinical groups (at health facility level)** |  |
| 21 | Patients initiating anti-TNF treatment |
| 22 | Patients receiving dialysis |
| 23 | Patients preparing for an organ or hematological transplant |
| 24 | Persons with Silicosis |
| **Provider Initiated groups, other High-risk groups (at community level)** |  |
| 25 | Prisoners |
| 26 | Miners (exposed to silica dust) |
| 27 | People with risk factors for TB seeking health care (for e.g., diabetes) |
| 28 | Populations with structural risk factors for TB |
| 29 | General population in settings with ≥ 0.5% general prevalence |

##### **General structure of the Target Population model**

The Target Population calculations are facilitated by a general structure that uses a combination of data or expert inputs to define:

- Population size
  - The calculation of populations sizes is directly based on inputs for provider-initiate programs:
    - For HH contacts and PLHIV on ART this information is obtained from calculation and from the Spectrum AIM model [1] respectively.
    - For High Risk (HR) groups the user must define population sizes, for example the size of a prison population (as a percentage of the overall 15+ population) or of a displaced population to be screened for TB disease.
    - The calculation of population sizes for patient-initiated programs is detailed below.
- Prevalence estimates for active (for the treatment branch) and latent TB (for the prevention branch)
  - For HH contacts and PLHIV on ART default estimates for the prevalence of active and latent TB are provided [2] [1].
  - For HR groups the user defines active TB prevalence via a relative risk (RR) variable which expresses TB prevalence in a HR group relative to the general population.
- A screening and clinical assessment algorithm
  - TB Global Plan recommended algorithms are used for screening, as detailed below.
  - Each screening algorithm, comprising one or two screening steps, is associated with an overall sensitivity and specificity which are used in the estimation of true and false positive cases referred for diagnoses from the screening step.
- A diagnostic algorithm
  - While the mechanism allows for the specification of the proportion of cases referred to diagnostic evaluation that are evaluated with different methods, e.g., smear microscopy and rapid molecular test, we use the TB Global Plan’s specification of universal rapid molecular tests. The sensitivity and specificity of the methods are again used for estimating the number of true and false positive cases who are diagnosed and linked to treatment.

#### **The TB care cascade, additional details**

##### *TB Preventive Treatment (TPT)*

After the number of patients with a positive diagnostic evaluation in provider-initiated programs is estimated, the remaining TPT-naive population is eligible for TPT, either:

- Presumptively due to being at very high risk for TB disease, e.g., child contacts or PLHIV with low CD4 count, or
- Following testing for latent TB infection

The patient-initiated model structure does not capture any further detail beyond the size of the screened population (i.e. no assumptions are made about the prevalence of TB infection in these populations). Therefore, there is no TPT modeled in the patient-initiated groups, and we limit TB prevention (TPT) to provider-initiated programs – i.e. HH contacts, PLHIV on ART and HR groups.

The proportion of LTBI diagnosis done with each TB infection testing method (tuberculin skin test (TST) or interferon-gamma release assay (IGRA), is configured to the specifications of the TB Global Plan.

##### *Drug resistance testing and treatment*

The total number of patients diagnosed and initiated on treatment for TB disease is determined by aggregating patient types over all the target population variations by age and pulmonary status. The group that a notified patient comes from: patient initiated, household contacts, PLHIV on ART or high-risk groups, plays no role in the selection of appropriate regimen used for treatment.

Regimen volumes recommended for defined patient groups are based on resistance profiles, which amount to specifying the proportion of patients that are:

- Rifampicin sensitive, with Isoniazid sensitive or Isoniazid resistant subsets
- Rifampicin resistant, with subsets that are Fluoroquinolone sensitive, Fluoroquinolone resistant (i.e. Pre-XDR) and XDR (Pre-XDR and resistant to at least one of Bedaquiline, Linezolid, Levofloacin, Moxifloxacin)

This resistance profile can only be established with full coverage of drug sensitivity testing:

- Rifampicin or simultaneous Rifampicin and Isoniazid resistance testing among patients with unknown Rifampicin and Isoniazid resistance
- Isoniazid resistance testing, among Rifampicin sensitive patients
- Fluoroquinolone resistance testing, among bacteriologically confirmed Rifampicin resistant patients
- Testing for resistance to Bedaquiline, Linezolid, Levofloxacin, Moxifloxacin (Pre-XDR or XDR)
- Patients who receive no DST receive default treatment for Rifampicin and Isoniazid sensitive patients.
- Rifampicin resistant patients receiving no DST for fluoroquinolone resistance receive the same regimen as those with confirmed fluoroquinolone sensitivity.

#### Algorithms for patient-initiated programs

**Pulmonary TB**

Description: For all persons attending health facilities with TB symptoms, the model assumes that all of them will be offered a WHO recommended molecular rapid diagnostic test. Those who test Mtb positive will be further offered other DR-TB tests such Xpert MTB/XDR test or liquid culture test with first- and second-line DST or targeted genome sequencing test (when it becomes available) to identify the resistance pattern and to assess the choice of anti-TB treatment regimen. Based on the results of the diagnostic tests, patients will be classified into the following five categories. Patients without resistance to HR, patients with H resistance (mono or poly) without resistance to rifampicin, patients with R or HR resistance only, patients with HR+FQ resistance and finally patients with HR+FQ+BDQ or injectable resistance and will be offered TB treatment regimen accordingly.

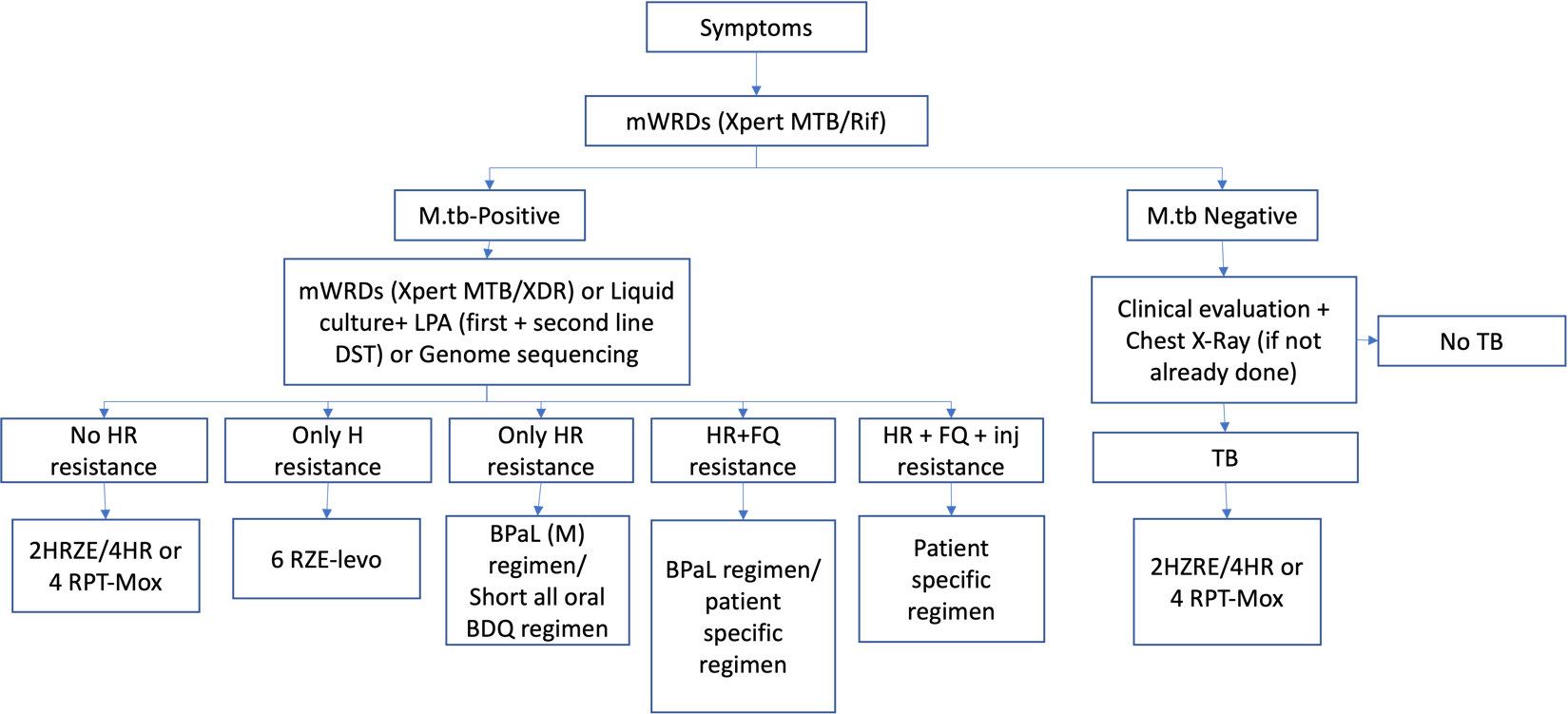

**Figure 1:** Algorithm-1 for diagnosis and treatment of Pulmonary TB in persons presenting with TB symptoms at health facilities (patient initiated)

The distribution of patients to various TB treatment regimens is based on the following assumptions given in Figure 2

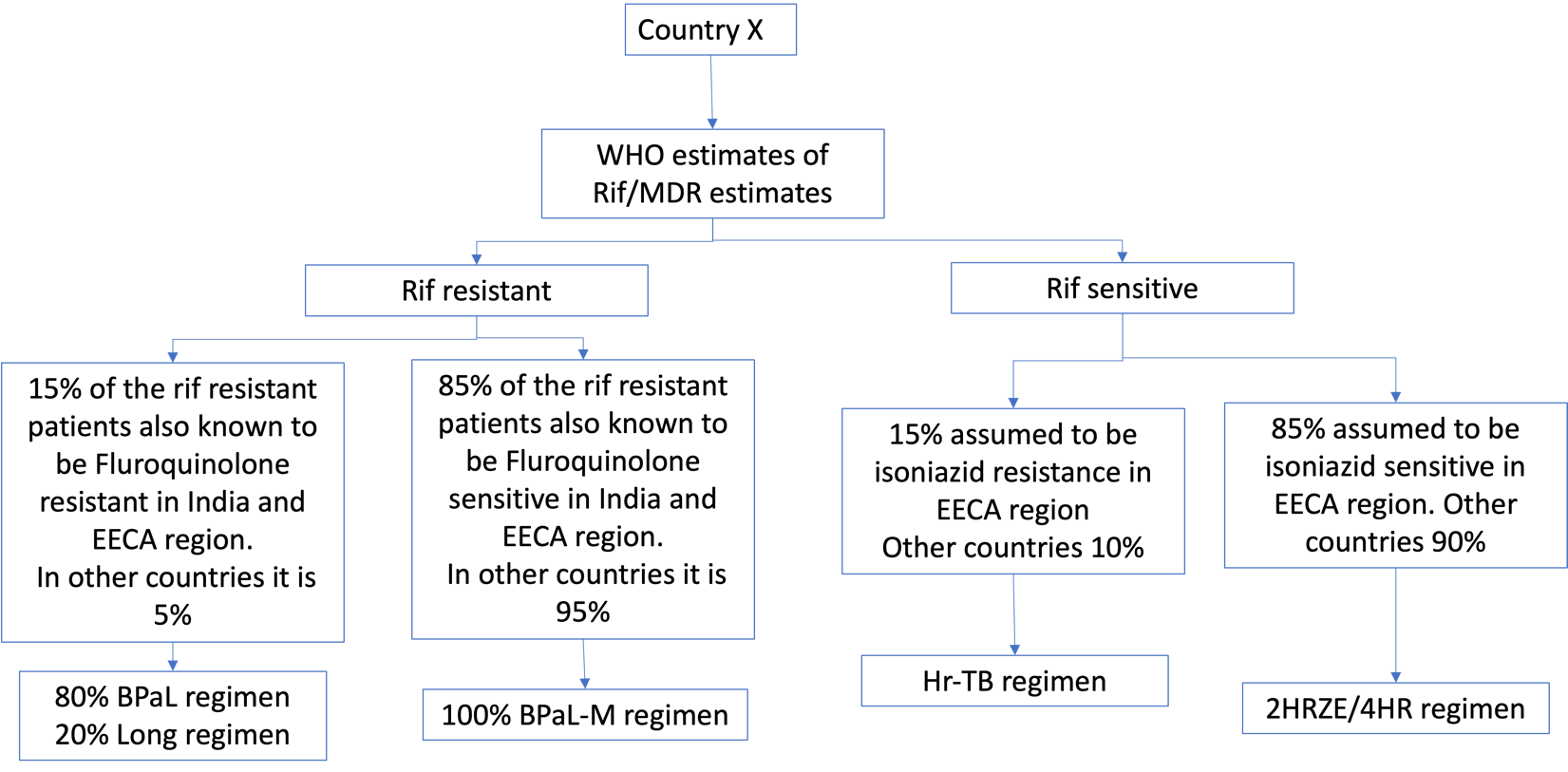

**Figure 2:** The methodology for estimating the proportion of TB patients eligible for various TB treatment regimens is as follows. Note that the assumed proportions can be varied by region or country, although for global modeling purposes we expect to use one set of assumed/estimated proportions.

In any given country, information about the prevalence of RR/ MDR-TB was obtained from the WHO’s Global TB report 2023. All patients with H and R sensitive TB (irrespective of resistance to other drugs) would receive the standard 2HRZE/4 HR TB treatment regimen. In patients with rifampicin sensitive TB, 15% in ECCA region and 10% in other countries were presumed to have Isoniazid mono or poly resistance and, in such patients, Hr-TB regimen will be provided. In those with RR/MDR-TB, 15% of such patients in India and EECA were assumed to be having fluroquinolone resistance and in other countries 5% of the patients were assumed to be having fluroquinolone resistance. 80% of the patients with fluroquinolone resistance would receive BPAL regimen and the remaining patients would receive longer DR-TB treatment regimen. In those with rifampicin resistant TB without fluroquinolone resistance, 100% of them would receive a BPaL-M regimen.

**Extra-pulmonary TB**

Description: For patients attending health facilities with extra-pulmonary TB symptoms, the model assumes that all of them will be offered a Chest Xray+ CAD, and a certain proportion will be additionally offered one or more of the following: CT-Scan, Ultrasound, FNAC Biopsy. This will be followed by a WHO recommended molecular rapid diagnostic test (Children would additionally receive a gastric lavage and PLHIV would receive a Urine LAM test). Those who test Mtb positive will be further offered other DR-TB tests such Xpert MTB/XDR test or liquid culture test with first- and second-line DST or targeted genome sequencing test (when it becomes available) to identify the resistance pattern and to assess the choice of anti-TB treatment regimen. Those who have a Mtb negative test will undergo clinical evaluation to determine if they have TB. If they have TB, they will receive an anti-TB treatment regimen (mostly drug sensitive anti-TB regimen with a few patients receiving DR-TB treatment regimen).

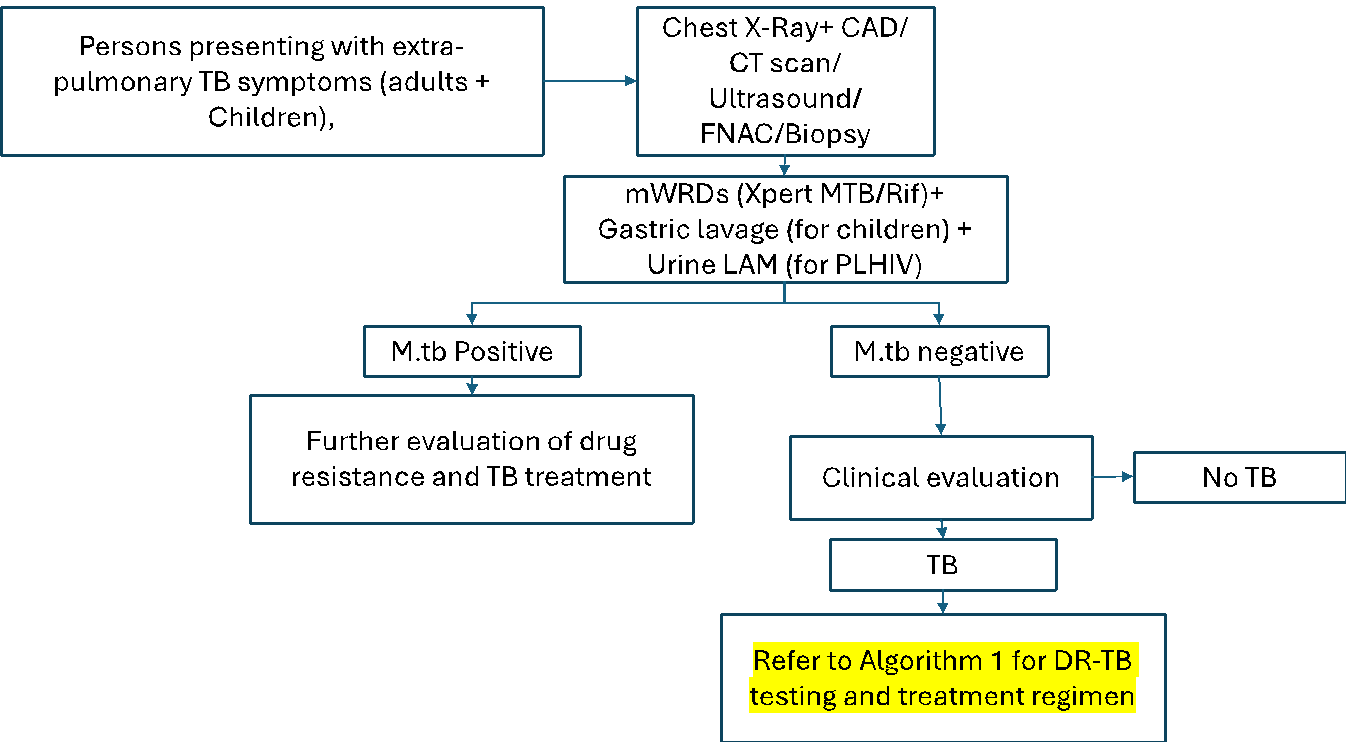

**Figure 3:** Algorithm for patients attending health facilities with extra-pulmonary TB symptoms.

#### **Algorithms for provider-initiated programs**

*Household contacts/ Contacts and other high risk clinical groups*

Description: For household contacts, the model assumes that all of them will be screened for TB symptoms and offered a Chest X-Ray with CAD (CAD for adults aged >15 years). Those who screen positive will be offered a mWRDT such as Xpert MTB/Rif and will follow the evaluation process as discussed for pulmonary (as shown in algorithm 1) and extra-pulmonary TB. In those without TB, TPT will be offered to everyone under the age of 5 years and to those aged > 5 years, TPT will be offered to those who test positive for TB infection (IGRA test).

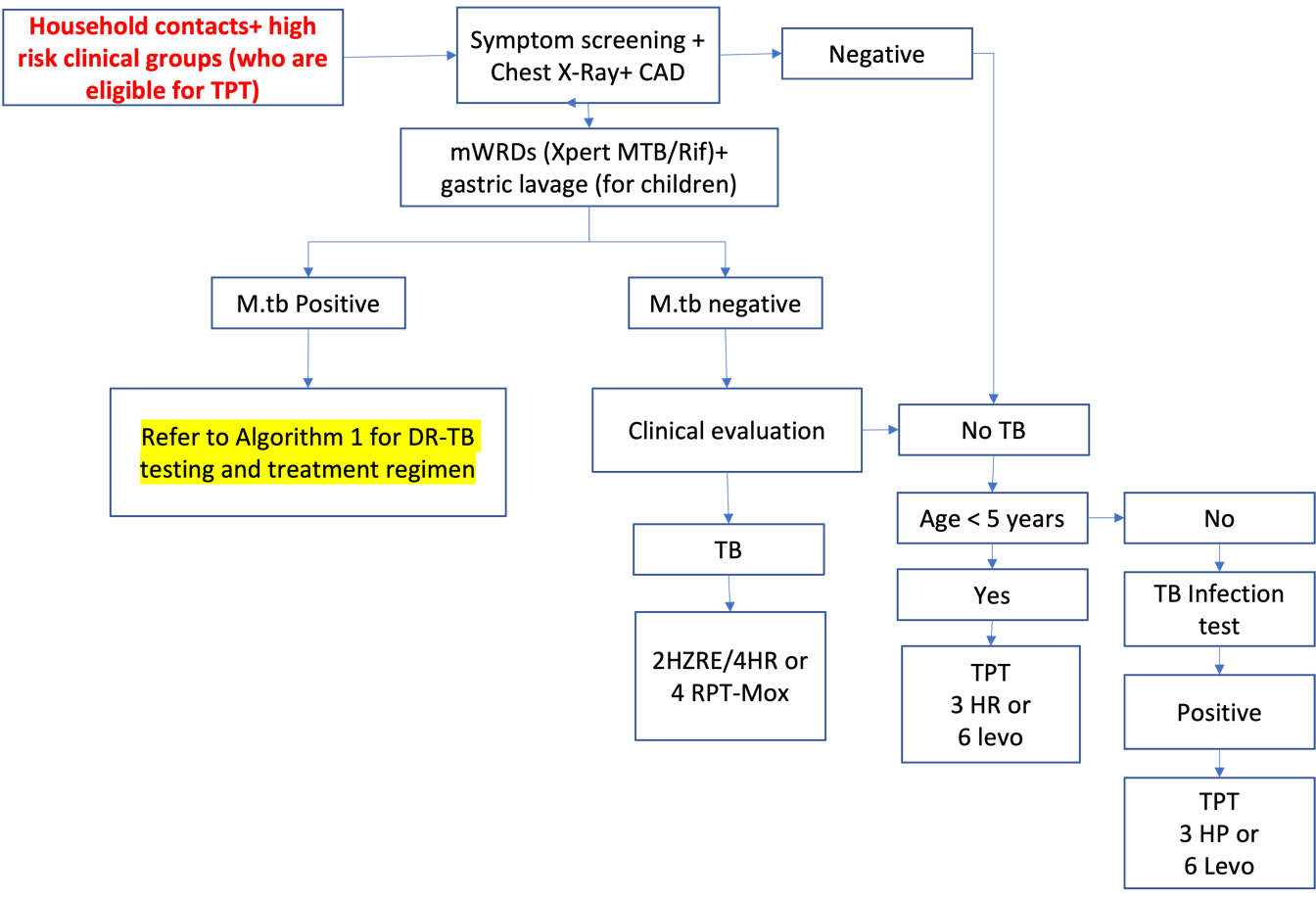

**Figure 4:** Algorithm-2 for TB case detection, assessing TPT eligibility and TPT provision among household contacts and other clinical high-risk groups

**People living with HIV**

Description

- For newly diagnosed with HIV, the model assumes that they would undergo screening for TB disease at the time of diagnosis. The screening methods includes 4 symptom screening along with a test for C-Reactive protein.
- For PLHIV who are already on ART, the model assumes that they would undergo systematic screening for TB disease once every year. The screening methods includes 4 symptom screening along with a Chest Radiography with CAD.
- Those who screen positive will be evaluated for pulmonary and extra-pulmonary TB disease using a combination of Urine LF-LAM test and mWRDT (Xpert MTB/Rif test) and will be treated for TB (if diagnosed). Newly diagnosed PLHIV without TB disease will be initiated on TPT.

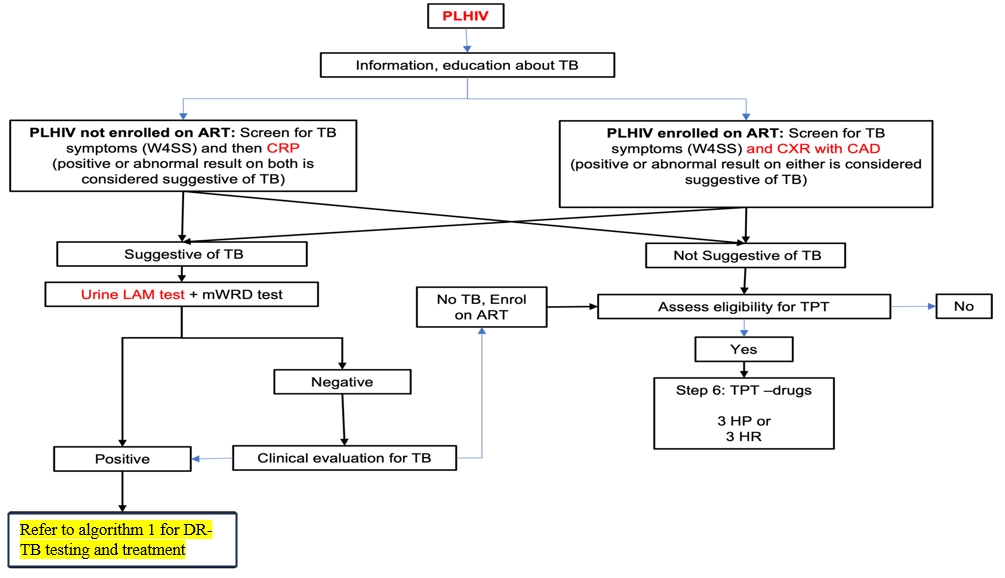

**Figure 5:** Algorithm-3 for systematic screening of PLHIV on ART for TB disease. Practically, screening and diagnosis for PLHIV not on ART is treated in the patient-initiated program, partly to avoid overlap between patient and provider-initiated programs with respect to PLHIV.

**Systematic screening in general population groups (at the community level)**

**Algorithm for systematic screening for TB (Figure 6)**

Description: All persons in the high-risk groups will be offered symptom screening and Chest X-Ray with CAD. Those who screen positive to either of the two screening methods will be offered mWRDs (Xpert MTB/Rif) test.

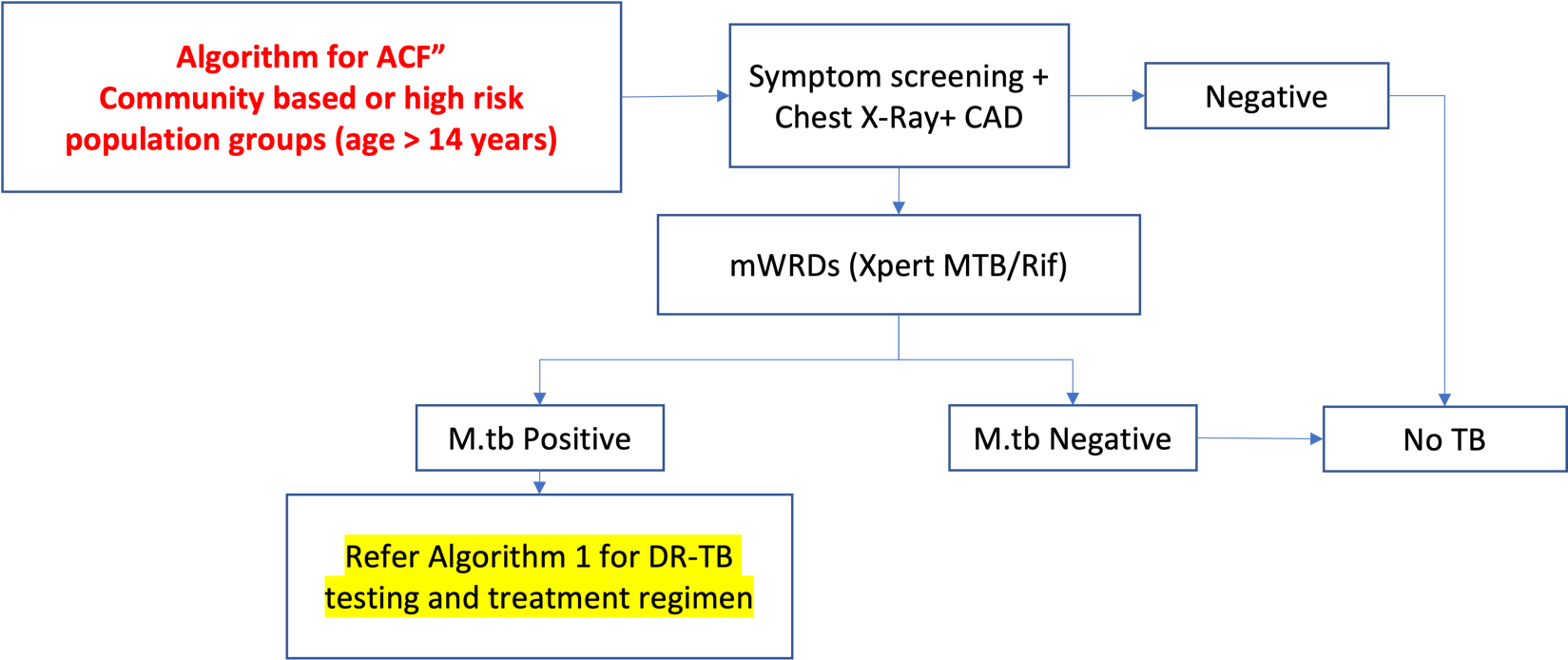

**Figure 6:** Algorithm-5 for systematic screening of high-risk groups for TB

#### Quantity and Population in Need (PIN) of screening and diagnostic interventions/service

*Table 3: For diagnosis: Patient-initiated Pulmonary TB among children aged <15 years, the following services, quantities and population in-need (PIN) are included for costing.*

| **Intervention/service** | **Quantity** | Pulmonary TB: HIV-negative, Children aged < 15 years (PIN) | Pulmonary TB: PLHIV not on ART, Children aged 0 to 9 years (PIN) | Pulmonary TB: PLHIV not on ART, Children aged 10 to 14 years (PIN) |
| --- | --- | --- | --- | --- |
| **Screening and Diagnosis** |  |  |  |  |
| screening visit to health facility | 1 | 100% | 100% | 100% |
| diagnostic visit to health facility | 1 | 100% | 100% | 100% |
| sputum/specimen collection and transportation | 1 | 40% | 40% | 40% |
| chest X-ray | 1 | 90% | 90% | 90% |
| computer-aided detection | 1 | 90% | 90% | 90% |
| portable-chest X-ray | 1 | 0% | 0% | 0% |
| sputum smear microscopy | 1 | 100% | 100% | 100% |
| gastric aspiration (for collecting specimens for diagnosis of TB in children) | 1 | 30% | 50% | 0% |
| lateral flow urine lipoarabinomannan assay | 1 | 0% | 20% | 20% |
| molecular WHO-recommended rapid diagnostic test | 1 | 40% | 40% | 40% |
| molecular WHO-recommended rapid diagnostic test for detection of rifampicin resistance | 1 | 40% | 40% | 40% |
| molecular WHO-recommended rapid diagnostic test for detection of rifampicin resistance | 1 | 60% | 60% | 60% |
| molecular WHO-recommended rapid diagnostic test for detection of resistance to first and second-line drugs | 1 | 100% | 100% | 100% |
| liquid culture | 1 | 100% | 100% | 100% |
| community visit | 1 | 0% | 0% | 0% |
| line probe assay for first-line drug | 1 | 5% | 5% | 5% |
| line probe assay for second-line drug | 1 | 5% | 5% | 5% |
| c-reactive protein | 1 | 0% | 15% | 15% |
| targeted gene sequencing | 1 | 0% | 0% | 0% |
| fine needle aspiration cytology | 1 | 0% | 0% | 0% |
| computed tomography scan | 1 | 0% | 0% | 0% |
| contact tracing visit | 1 | 0% | 0% | 0% |
| tuberculin skin test | 1 | 0% | 0% | 0% |
| interferon-gamma release assay | 1 | 0% | 0% | 0% |
| TB antigen-based skin tests (TBSTs) | 1 | 0% | 0% | 0% |
| biopsy | 1 | 0% | 0% | 0% |
| ultrasonography | 1 | 0% | 0% | 0% |
| magnetic resonance imaging | 1 | 0% | 0% | 0% |

*Table 4: For diagnosis: Patient-initiated extra-pulmonary among children aged <15 years, the following services, quantities and population in-need (PIN) are included for costing*

| **Intervention** | **Quantity** | Extra-Pulmonary TB: HIV-negative, Children aged < 15 years (PIN) | Extra-Pulmonary TB: PLHIV not on ART, Children aged 0 to 9 years (PIN | Extra-Pulmonary TB: PLHIV on ART, Children aged 10 to 14 years (PIN) |
| --- | --- | --- | --- | --- |
| **Screening and Diagnosis** |  |  |  |  |
| screening visit to health facility | 1 | 100% | 100% | 100% |
| diagnostic visit to health facility | 1 | 100% | 100% | 100% |
| sputum/specimen collection and transportation | 1 | 50% | 50% | 50% |
| chest X-ray | 1 | 100% | 100% | 100% |
| computer-aided detection | 1 | 100% | 100% | 100% |
| portable-chest X-ray | 1 | 0% | 0% | 0% |
| sputum smear microscopy | 1 | 100% | 100% | 100% |
| gastric aspiration (for collecting specimens for diagnosis of TB in children) | 1 | 30% | 50% | 0% |
| lateral flow urine lipoarabinomannan assay | 1 | 0% | 20% | 20% |
| molecular WHO-recommended rapid diagnostic test | 1 | 40% | 40% | 40% |
| molecular WHO-recommended rapid diagnostic test for detection of rifampicin resistance | 1 | 40% | 40% | 40% |
| molecular WHO-recommended rapid diagnostic test for detection of rifampicin resistance | 1 | 60% | 60% | 60% |
| molecular WHO-recommended rapid diagnostic test for detection of resistance to first and second-line drugs | 1 | 100% | 100% | 100% |
| liquid culture | 1 | 100% | 100% | 100% |
| community visit | 1 | 0% | 0% | 0% |
| line probe assay for first-line drug | 1 | 5% | 5% | 5% |
| line probe assay for second-line drug | 1 | 5% | 5% | 5% |
| c-reactive protein | 1 | 0% | 15% | 15% |
| targeted gene sequencing | 1 | 0% | 0% | 0% |
| fine needle aspiration cytology | 1 | 60% | 60% | 60% |
| computed tomography scan | 1 | 20% | 20% | 20% |
| contact tracing visit | 1 | 0% | 0% | 0% |
| tuberculin skin test | 1 | 0% | 0% | 0% |
| interferon-gamma release assay | 1 | 0% | 0% | 0% |
| TB antigen-based skin tests (TBSTs) | 1 | 0% | 0% | 0% |
| biopsy | 1 | 60% | 60% | 60% |
| ultrasonography | 1 | 50% | 50% | 50% |
| magnetic resonance imaging | 1 | 20% | 20% | 20% |

*Table 5: For diagnosis: Patient-initiated, Pulmonary adults (age >15 years)*

| **Intervention** | **Quantity** | Pulmonary TB: HIV-negative, Adults 15 years and above (PIN) | Pulmonary TB: PLHIV not on ART, Adults aged 15 years and above (PIN) |
| --- | --- | --- | --- |
| **Screening and Diagnosis** |  |  |  |
| screening visit to health facility | 1 | 100% | 100% |
| diagnostic visit to health facility | 1 | 100% | 100% |
| sputum/specimen collection and transportation | 1 | 40% | 40% |
| chest X-ray | 1 | 90% | 20% |
| computer-aided detection | 1 | 90% | 20% |
| portable-chest X-ray | 1 | 0% | 0% |
| sputum smear microscopy | 1 | 100% | 100% |
| gastric aspiration (for collecting specimens for diagnosis of TB in children) | 1 | 0% | 0% |
| lateral flow urine lipoarabinomannan assay | 1 | 0% | 20% |
| molecular WHO-recommended rapid diagnostic test | 1 | 40% | 40% |
| molecular WHO-recommended rapid diagnostic test for detection of rifampicin resistance | 1 | 40% | 40% |
| molecular WHO-recommended rapid diagnostic test for detection of rifampicin resistance | 1 | 60% | 60% |
| molecular WHO-recommended rapid diagnostic test for detection of resistance to first and second-line drugs | 1 | 100% | 100% |
| liquid culture | 1 | 100% | 100% |
| community visit | 1 | 0% | 0% |
| line probe assay for first-line drug | 1 | 5% | 5% |
| line probe assay for second-line drug | 1 | 5% | 5% |
| c-reactive protein | 1 | 0% | 15% |
| targeted gene sequencing | 1 | 0% | 0% |
| fine needle aspiration cytology | 1 | 0% | 0% |
| computed tomography scan | 1 | 0% | 0% |
| contact tracing visit | 1 | 0% | 0% |
| tuberculin skin test | 1 | 0% | 0% |
| interferon-gamma release assay | 1 | 0% | 0% |
| TB antigen-based skin tests (TBSTs) | 1 | 0% | 0% |
| biopsy | 1 | 0% | 0% |
| ultrasonography | 1 | 0% | 0% |
| magnetic resonance imaging | 1 | 0% | 0% |

*Table 6: For diagnosis: Patient-initiated extra-pulmonary (age>15 years)*

| **Intervention** | **Quantity** | Extra-Pulmonary TB: HIV-negative, Adults 15 years and above (PIN) | Extra-Pulmonary TB: PLHIV not on ART, Adults aged 15 years and above (PIN) |
| --- | --- | --- | --- |
| **Screening and Diagnosis** |  |  |  |
| screening visit to health facility | 1 | 100% | 100% |
| diagnostic visit to health facility | 1 | 100% | 100% |
| sputum/specimen collection and transportation | 1 | 50% | 50% |
| chest X-ray | 1 | 100% | 50% |
| computer-aided detection | 1 | 100% | 50% |
| portable-chest X-ray | 1 | 0% | 0% |
| sputum smear microscopy | 1 | 100% | 100% |
| gastric aspiration (for collecting specimens for diagnosis of TB in children) | 1 | 0% | 0% |
| lateral flow urine lipoarabinomannan assay | 1 | 0% | 20% |
| molecular WHO-recommended rapid diagnostic test | 1 | 40% | 40% |
| molecular WHO-recommended rapid diagnostic test for detection of rifampicin resistance | 1 | 40% | 40% |
| molecular WHO-recommended rapid diagnostic test for detection of rifampicin resistance | 1 | 60% | 60% |
| molecular WHO-recommended rapid diagnostic test for detection of resistance to first and second-line drugs | 1 | 100% | 100% |
| liquid culture | 1 | 100% | 100% |
| community visit | 1 | 0% | 0% |
| line probe assay for first-line drug | 1 | 5% | 5% |
| line probe assay for second-line drug | 1 | 5% | 5% |
| c-reactive protein | 1 | 0% | 15% |
| targeted gene sequencing | 1 | 0% | 0% |
| fine needle aspiration cytology | 1 | 60% | 60% |
| computed tomography scan | 1 | 20% | 20% |
| contact tracing visit | 1 | 0% | 0% |
| tuberculin skin test | 1 | 0% | 0% |
| interferon-gamma release assay | 1 | 0% | 0% |
| TB antigen-based skin tests (TBSTs) | 1 | 0% | 0% |
| biopsy | 1 | 60% | 60% |
| ultrasonography | 1 | 50% | 50% |
| magnetic resonance imaging | 1 | 20% | 20% |

*Table 7: For diagnosis: Provider-initiated, Household contacts*

| **Intervention** | **Quantity** | HH Contacts, Pulmonary TB: HIV-negative, Children aged 0 to 4 years (PIN) | HH Contacts, Pulmonary TB: HIV-negative, Children aged 5 to 14 years (PIN) | HH Contacts, Pulmonary TB: HIV-negative, Children aged 15 years and above (PIN) |
| --- | --- | --- | --- | --- |
| **Screening and Diagnosis** |  |  |  |  |
| screening visit to health facility | 1 | 100% | 100% | 100% |
| diagnostic visit to health facility | 1 | 10% | 10% | 5% |
| sputum/specimen collection and transportation | 1 | 75% | 75% | 10% |
| chest X-ray | 1 | 90% | 90% | 90% |
| computer-aided detection | 1 | 90% | 90% | 90% |
| portable-chest X-ray | 1 | 0% | 0% | 0% |
| sputum smear microscopy | 1 | 100% | 100% | 100% |
| gastric aspiration (for collecting specimens for diagnosis of TB in children) | 1 | 40% | 0% | 0% |
| lateral flow urine lipoarabinomannan assay | 1 | 0% | 0% | 0% |
| molecular WHO-recommended rapid diagnostic test | 1 | 40% | 40% | 40% |
| molecular WHO-recommended rapid diagnostic test for detection of rifampicin resistance | 1 | 40% | 40% | 40% |
| molecular WHO-recommended rapid diagnostic test for detection of rifampicin resistance | 1 | 60% | 60% | 60% |
| molecular WHO-recommended rapid diagnostic test for detection of resistance to first- and second-line drugs | 1 | 100% | 100% | 100% |
| liquid culture | 1 | 100% | 100% | 100% |
| community visit | 1 | 0% | 0% | 0% |
| line probe assay for first-line drug | 1 | 5% | 5% | 5% |
| line probe assay for second-line drug | 1 | 5% | 5% | 5% |
| c-reactive protein | 1 | 0% | 0% | 0% |
| targeted gene sequencing | 1 | 0% | 0% | 0% |
| fine needle aspiration cytology | 1 | 15% | 15% | 5% |
| computed tomography scan | 1 | 5% | 5% | 1% |
| contact tracing visit | 1 | 100% | 100% | 100% |
| tuberculin skin test | 1 | 0% | 10% | 10% |
| interferon-gamma release assay | 1 | 0% | 90% | 90% |
| TB antigen-based skin tests (TBSTs) | 1 | 0% | 0% | 0% |
| biopsy | 1 | 15% | 15% | 5% |
| ultrasonography | 1 | 15% | 15% | 1% |
| magnetic resonance imaging | 1 | 2% | 2% | 0% |

*Table 8: For diagnosis: Provider-initiated, CLHIV*

| **Intervention** | **Quantity** | CLHIV, Pulmonary TB: PLHIV on ART with severe disease, Children aged 0 to 9 years (PIN) | CLHIV, Pulmonary TB: PLHIV on ART without severe disease, Children aged 0 to 9 years (PIN) | CLHIV, Pulmonary TB: PLHIV on ART with severe disease, Children aged 10 to 14 years (PIN) | CLHIV, Pulmonary TB: PLHIV on ART without severe disease, Children aged 10 to 14 years (PIN) |
| --- | --- | --- | --- | --- | --- |
| **Screening and Diagnosis** |  |  |  |  |  |
| screening visit to health facility | 1 | 100% | 100% | 100% | 100% |
| diagnostic visit to health facility | 1 | 40% | 40% | 40% | 40% |
| sputum/specimen collection and transportation | 1 | 50% | 20% | 50% | 20% |
| chest X-ray | 1 | 90% | 90% | 90% | 90% |
| computer-aided detection | 1 | 90% | 100% | 90% | 100% |
| portable-chest X-ray | 1 | 0% | 0% | 0% | 0% |
| sputum smear microscopy | 1 | 100% | 100% | 100% | 100% |
| gastric aspiration (for collecting specimens for diagnosis of TB in children) | 1 | 50% | 40% | 0% | 0% |
| lateral flow urine lipoarabinomannan assay | 1 | 20% | 20% | 20% | 20% |
| molecular WHO-recommended rapid diagnostic test | 1 | 40% | 40% | 40% | 40% |
| molecular WHO-recommended rapid diagnostic test for detection of rifampicin resistance | 1 | 40% | 40% | 40% | 40% |
| molecular WHO-recommended rapid diagnostic test for detection of rifampicin resistance | 1 | 60% | 60% | 60% | 60% |
| molecular WHO-recommended rapid diagnostic test for detection of resistance to first and second-line drugs | 1 | 100% | 100% | 100% | 100% |
| liquid culture | 1 | 100% | 100% | 100% | 100% |
| community visit | 1 | 0% | 0% | 0% | 0% |
| line probe assay for first-line drug | 1 | 5% | 5% | 5% | 5% |
| line probe assay for second-line drug | 1 | 5% | 5% | 5% | 5% |
| c-reactive protein | 1 | 15% | 15% | 15% | 15% |
| targeted gene sequencing | 1 | 0% | 0% | 0% | 0% |
| fine needle aspiration cytology | 1 | 40% | 10% | 40% | 10% |
| computed tomography scan | 1 | 10% | 1% | 10% | 1% |
| contact tracing visit | 1 | 0% | 0% | 0% | 0% |
| tuberculin skin test | 1 | 0% | 0% | 0% | 0% |
| interferon-gamma release assay | 1 | 0% | 0% | 0% | 0% |
| TB antigen-based skin tests (TBSTs) | 1 | 0% | 0% | 0% | 0% |
| biopsy | 1 | 40% | 10% | 40% | 10% |
| ultrasonography | 1 | 40% | 10% | 40% | 10% |
| magnetic resonance imaging | 1 | 5% | 1% | 5% | 1% |

*Table 9: For diagnosis: Provider-initiated, PLHIV adults >15 years*

| **Intervention** | **Quantity** | PLHIV, Pulmonary TB: PLHIV on ART with severe disease, Adults aged 15 years and above (PIN) | PLHIV, Pulmonary TB: PLHIV on ART without severe disease, Adults aged 15 years and above (PIN) |
| --- | --- | --- | --- |
| **Screening and Diagnosis** |  |  |  |
| screening visit to health facility | 1 | 100% | 100% |
| diagnostic visit to health facility | 1 | 100% | 40% |
| sputum/specimen collection and transportation | 1 | 20% | 20% |
| chest X-ray | 1 | 90% | 90% |
| computer-aided detection | 1 | 90% | 90% |
| portable-chest X-ray | 1 | 0% | 0% |
| sputum smear microscopy | 1 | 100% | 100% |
| gastric aspiration (for collecting specimens for diagnosis of TB in children) | 1 | 0% | 0% |
| lateral flow urine lipoarabinomannan assay | 1 | 20% | 20% |
| molecular WHO-recommended rapid diagnostic test | 1 | 40% | 40% |
| molecular WHO-recommended rapid diagnostic test for detection of rifampicin resistance | 1 | 40% | 40% |
| molecular WHO-recommended rapid diagnostic test for detection of rifampicin resistance | 1 | 60% | 60% |
| molecular WHO-recommended rapid diagnostic test for detection of resistance to first and second line drugs | 1 | 100% | 100% |
| liquid culture | 1 | 100% | 100% |
| community visit | 1 | 0% | 0% |
| line probe assay for first-line drug | 1 | 5% | 5% |
| line probe assay for second-line drug | 1 | 5% | 5% |
| c-reactive protein | 1 | 15% | 15% |
| targeted gene sequencing | 1 | 0% | 0% |
| fine needle aspiration cytology | 1 | 25% | 10% |
| computed tomography scan | 1 | 25% | 5% |
| contact tracing visit | 1 | 0% | 0% |
| tuberculin skin test | 1 | 0% | 0% |
| interferon-gamma release assay | 1 | 0% | 0% |
| TB antigen-based skin tests (TBSTs) | 1 | 0% | 0% |
| biopsy | 1 | 10% | 10% |
| ultrasonography | 1 | 25% | 10% |
| magnetic resonance imaging | 1 | 10% | 0% |

*Table 10: For diagnosis: Provider-initiated, high-risk groups*

| **Intervention** | **Quantity** | HR groups, Pulmonary TB: HIV-negative, Adults aged 15 years and above (PIN) |
| --- | --- | --- |
| **Screening and Diagnosis** |  |  |
| screening visit to health facility | 1 | 100.0% |
| diagnostic visit to health facility | 1 | 5.0% |
| sputum/specimen collection and transportation | 1 | 2.5% |
| chest X-ray | 1 | 0.0% |
| computer-aided detection | 1 | 90.0% |
| portable-chest X-ray | 1 | 90.0% |
| sputum smear microscopy | 1 | 100.0% |
| gastric aspiration (for collecting specimens for diagnosis of TB in children) | 1 | 0.0% |
| lateral flow urine lipoarabinomannan assay | 1 | 0.0% |
| molecular WHO-recommended rapid diagnostic test | 1 | 40.0% |
| molecular WHO-recommended rapid diagnostic test for detection of rifampicin resistance | 1 | 40.0% |
| molecular WHO-recommended rapid diagnostic test for detection of rifampicin resistance | 1 | 60.0% |
| molecular WHO-recommended rapid diagnostic test for detection of resistance to first and second-line drugs | 1 | 100.0% |
| liquid culture | 1 | 100.0% |
| community visit | 1 | 100.0% |
| line probe assay for first-line drug | 1 | 5.0% |
| line probe assay for second-line drug | 1 | 5.0% |
| c-reactive protein | 1 | 0.0% |
| targeted gene sequencing | 1 | 0.0% |
| fine needle aspiration cytology | 1 | 0.0% |
| computed tomography scan | 1 | 0.0% |
| contact tracing visit | 1 | 0.0% |
| tuberculin skin test | 1 | 10.0% |
| interferon-gamma release assay | 1 | 90.0% |
| TB antigen-based skin tests (TBSTs) | 1 | 0.0% |
| biopsy | 1 | 0.0% |
| ultrasonography | 1 | 0.0% |
| magnetic resonance imaging | 1 | 0.0% |

#### Quantity and Population in Need (PIN) of treatment interventions/service

*Table 11: Pediatric TB treatment: Interventions/services (quantities, in accordance with WHO guidelines)*

| **Intervention/ services** | **2HRZE/**  **4HR** | **6HRZEto** | **4-month shorter regimen [2HRZ(E)/2HR]** | **Regimen for Isoniazid resistant TB (Hr-TB )** | **BPaL M** | **Short all-oral BDQ** | **BPAL** | **Delamanid-based regimen longer regimen** |
| --- | --- | --- | --- | --- | --- | --- | --- | --- |
| patient counselling | 2 | 2 | 2 | 2 | 2 | 2 | 2 | 2 |
| treatment monitoring | 6 | 6 | 4 | 6 | 6 | 9 | 6 | 18 |
| outpatient department treatment visit | 3 | 6 | 2 | 3 | 6 | 9 | 6 | 18 |
| In patient management of Drug sensitive TB | 5 | 10 | 5 | 0 | 0 | 0 | 0 | 0 |
| In patient management of Drug-resistant TB | 0 | 0 | 0 | 5 | 5 | 5 | 5 | 10 |
| directly observed treatment | 1 | 1 | 1 | 1 | 1 | 1 | 1 | 1 |
| digital adherence technology | 1 | 1 | 1 | 1 | 1 | 1 | 1 | 1 |
| gastric aspiration (for collecting specimens for diagnosis of TB in children) | 1 | 1 | 1 | 1 | 1 | 1 | 1 | 1 |
| sputum smear microscopy | 2 | 2 | 2 | 2 | 6 | 9 | 6 | 18 |
| sputum culture monthly | 0 | 0 | 0 | 1 | 6 | 9 | 6 | 18 |
| electrocardiogram | 1 | 1 | 1 | 1 | 3 | 3 | 3 | 4 |
| liver function test | 1 | 1 | 1 | 1 | 1 | 1 | 1 | 4 |
| diabetes mellitus | 1 | 1 | 1 | 1 | 1 | 1 | 1 | 1 |
| renal function test | 1 | 1 | 1 | 1 | 2 | 2 | 2 | 4 |
| serum glutamic pyruvic transaminase | 1 | 1 | 1 | 1 | 2 | 2 | 2 | 6 |
| serum glutamic-oxaloacetic transaminase | 1 | 1 | 1 | 1 | 2 | 2 | 2 | 6 |
| ultrasonography | 1 | 2 | 1 | 1 | 1 | 1 | 1 | 1 |
| contact tracing visit | 1 | 1 | 1 | 1 | 1 | 1 | 1 | 1 |
| magnetic resonance imaging | 1 | 1 | 1 | 1 | 1 | 1 | 1 | 1 |
| tracing those who are lost to follow-up | 1 | 1 | 1 | 1 | 1 | 1 | 1 | 1 |
| patient support costs | 1 | 1 | 1 | 1 | 1 | 1 | 1 | 1 |
| active TB drug safety monitoring and management | 1 | 1 | 1 | 1 | 1 | 1 | 1 | 1 |
| adverse event management through IP care | 1 | 1 | 1 | 1 | 1 | 1 | 1 | 1 |
| elective partial lung resection (lobectomy or wedge resection) | 1 | 1 | 1 | 1 | 1 | 1 | 1 | 1 |
| follow-up post-TB treatment | 1 | 3 | 1 | 1 | 1 | 1 | 1 | 1 |
| Post TB lung disease care | 1 | 1 | 1 | 1 | 1 | 1 | 1 | 1 |
| Palliative care | 1 | 1 | 1 | 1 | 1 | 1 | 1 | 1 |

*Table 12: Pediatric TB treatment: Interventions/services (PINs)*

| **Intervention** | **2HRZE/**  **4HR** | **6HRZEto** | **4-month shorter regimen [2HRZ(E)/2HR]** | **Regimen for Isoniazid resistant TB (Hr-TB )** | **BPaL M** | **Short all-oral BDQ** | **BPAL** | **Delamanid-based regimen longer regimen** |
| --- | --- | --- | --- | --- | --- | --- | --- | --- |
| patient counselling | 100% | 100% | 100% | 100% | 100% | 100% | 100% | 100% |
| treatment monitoring | 100% | 100% | 100% | 100% | 100% | 100% | 100% | 100% |
| outpatient department treatment visit | 100% | 100% | 100% | 100% | 100% | 100% | 100% | 100% |
| In patient management of Drug sensitive TB | 20% | 100% | 5% | 0% | 0% | 0% | 0% | 0% |
| In patient management of Drug-resistant TB | 0% | 0% | 0% | 20% | 20% | 20% | 20% | 50% |
| directly observed treatment | 0% | 0% | 0% | 0% | 0% | 0% | 0% | 0% |
| digital adherence technology | 100% | 100% | 100% | 100% | 100% | 100% | 100% | 100% |
| gastric aspiration (for collecting specimens for diagnosis of TB in children) | 20% | 20% | 20% | 20% | 20% | 20% | 20% | 20% |
| sputum smear microscopy | 95% | 95% | 95% | 95% | 95% | 95% | 95% | 95% |
| sputum culture monthly | 0% | 0% | 0% | 0% | 95% | 95% | 95% | 95% |
| electrocardiogram | 0% | 0% | 0% | 0% | 100% | 100% | 100% | 100% |
| liver function test | 10% | 10% | 10% | 100% | 100% | 100% | 100% | 100% |
| diabetes mellitus | 10% | 100% | 10% | 10% | 100% | 100% | 100% | 100% |
| renal function test | 10% | 100% | 10% | 10% | 100% | 100% | 100% | 100% |
| serum glutamic pyruvic transaminase | 10% | 100% | 10% | 10% | 20% | 20% | 20% | 100% |
| serum glutamic-oxaloacetic transaminase | 10% | 100% | 10% | 10% | 20% | 20% | 20% | 100% |
| ultrasonography | 20% | 100% | 0% | 20% | 20% | 20% | 20% | 100% |
| contact tracing visit | 0% | 0% | 0% | 0% | 0% | 0% | 0% | 0% |
| magnetic resonance imaging | 5% | 100% | 0% | 10% | 10% | 10% | 10% | 10% |
| tracing those who are lost to follow-up | 20% | 20% | 20% | 20% | 20% | 20% | 20% | 20% |
| patient support costs | 0% | 0% | 0% | 0% | 0% | 0% | 0% | 0% |
| active TB drug safety monitoring and management | 100% | 100% | 100% | 100% | 100% | 100% | 100% | 100% |
| adverse event management through IP care | 5% | 5% | 5% | 5% | 5% | 5% | 5% | 10% |
| elective partial lung resection (lobectomy or wedge resection) | 1% | 0% | 0% | 0% | 5% | 5% | 5% | 50% |
| follow-up post-TB treatment | 90% | 90% | 90% | 90% | 90% | 90% | 90% | 90% |
| Post TB lung disease care | 10% | 50% | 0% | 10% | 20% | 20% | 20% | 50% |
| Palliative care | 5% | 50% | 5% | 10% | 50% | 50% | 50% | 100% |

*Table 13:Adult TB treatment: Interventions/services (quantities)*

| **Intervention** | **2HRZE/**  **4HR** | **Four-month RPT-MOX regimen** | **Hr-TB regimen** | **BPaL M** | **Short all-oral BDQ regimen** | **BPaL** | **Long regimen for DR-TB, containing delamanid** |
| --- | --- | --- | --- | --- | --- | --- | --- |
| patient counselling | 2.0 | 2.0 | 2.0 | 2.0 | 2.0 | 2.0 | 2.0 |
| treatment monitoring | 6.0 | 4.0 | 6.0 | 6.0 | 9.0 | 6.0 | 18.0 |
| outpatient department treatment visit | 3.0 | 2.0 | 3.0 | 6.0 | 9.0 | 6.0 | 18.0 |
| In patient management of Drug sensitive TB | 5.0 | 5.0 | 5.0 | 0.0 | 0.0 | 0.0 | 0.0 |
| In patient management of Drug-resistant TB | 0.0 | 0.0 | 0.0 | 5.0 | 5.0 | 5.0 | 10.0 |
| directly observed treatment | 1.0 | 1.0 | 1.0 | 1.0 | 1.0 | 1.0 | 1.0 |
| digital adherence technology | 1.0 | 1.0 | 1.0 | 1.0 | 1.0 | 1.0 | 1.0 |
| gastric aspiration (for collecting specimens for diagnosis of TB in children) | 0.0 | 0.0 | 0.0 | 0.0 | 0.0 | 0.0 | 0.0 |
| sputum smear microscopy | 2.0 | 2.0 | 2.0 | 6.0 | 9.0 | 6.0 | 18.0 |
| sputum culture monthly | 0.0 | 0.0 | 0.0 | 6.0 | 9.0 | 6.0 | 18.0 |
| electrocardiogram | 1.0 | 1.0 | 1.0 | 3.0 | 3.0 | 3.0 | 4 |
| liver function test | 1.0 | 1.0 | 1.0 | 1.0 | 1.0 | 1.0 | 4 |
| diabetes mellitus | 1.0 | 1.0 | 1.0 | 1.0 | 1.0 | 1.0 | 1 |
| renal function test | 1.0 | 1.0 | 1.0 | 2.0 | 2.0 | 2.0 | 4 |
| serum glutamic pyruvic transaminase | 1.0 | 1.0 | 1.0 | 2.0 | 2.0 | 2.0 | 6 |
| serum glutamic-oxaloacetic transaminase | 1.0 | 1.0 | 1.0 | 2.0 | 2.0 | 2.0 | 6 |
| ultrasonography | 1.0 | 1.0 | 1.0 | 1.0 | 1.0 | 1.0 | 1.0 |
| contact tracing visit | 1.0 | 1.0 | 1.0 | 1.0 | 1.0 | 1.0 | 1.0 |
| magnetic resonance imaging | 1.0 | 1.0 | 1.0 | 1.0 | 1.0 | 1.0 | 1.0 |
| tracing those who are lost to follow-up | 1.0 | 1.0 | 1.0 | 1.0 | 1.0 | 1.0 | 1.0 |
| patient support costs | 1.0 | 1.0 | 1.0 | 1.0 | 1.0 | 1.0 | 1.0 |
| active TB drug safety monitoring and management | 1.0 | 1.0 | 1.0 | 1.0 | 1.0 | 1.0 | 1.0 |
| adverse event management through IP care | 1.0 | 1.0 | 1.0 | 1.0 | 1.0 | 1.0 | 1.0 |
| elective partial lung resection (lobectomy or wedge resection) | 1.0 | 1.0 | 1.0 | 1.0 | 1.0 | 1.0 | 1.0 |
| follow-up post-TB treatment | 1.0 | 1.0 | 1.0 | 1.0 | 1.0 | 1.0 | 1.0 |
| Post TB lung disease care | 1.0 | 1.0 | 1.0 | 1.0 | 1.0 | 1.0 | 1.0 |
| Palliative care | 1.0 | 1.0 | 1.0 | 1.0 | 1.0 | 1.0 | 1.0 |

*Table 14: Adult TB treatment: Interventions/services (PINs)*

| **Intervention** | **2HRZE/**  **4HR** | **Four-month RPT-MOX regimen** | **Hr-TB regimen** | **BPaL M** | **Short all-oral BDQ regimen** | **BPaL** | **Long regimen for DR-TB, containing delamanid** |
| --- | --- | --- | --- | --- | --- | --- | --- |
| patient counselling | 100% | 100% | 100% | 100% | 100% | 100% | 100% |
| treatment monitoring | 100% | 100% | 100% | 100% | 100% | 100% | 100% |
| outpatient department treatment visit | 100% | 100% | 100% | 100% | 100% | 100% | 100% |
| In patient management of Drug sensitive TB | 20% | 20% | 20% | 0% | 0% | 0% | 0% |
| In patient management of Drug resistant TB | 0% | 0% | 0% | 20% | 20% | 20% | 50% |
| directly observed treatment | 0% | 0% | 0% | 0% | 0% | 0% | 0% |
| digital adherence technology | 100% | 100% | 100% | 100% | 100% | 100% | 100% |
| gastric aspiration (for collecting specimens for diagnosis of TB in children) | 0% | 0% | 0% | 0% | 0% | 0% | 0% |
| sputum smear microscopy | 95% | 95% | 95% | 95% | 95% | 95% | 95% |
| sputum culture monthly | 0% | 0% | 0% | 95% | 95% | 95% | 95% |
| electrocardiogram | 0% | 0% | 0% | 100% | 100% | 100% | 100% |
| liver function test | 10% | 10% | 10% | 100% | 100% | 100% | 100% |
| diabetes mellitus | 1% | 10% | 10% | 100% | 100% | 100% | 100% |
| renal function test | 10% | 10% | 100% | 100% | 100% | 100% | 100% |
| serum glutamic pyruvic transaminase | 10% | 10% | 10% | 20% | 20% | 20% | 100% |
| serum glutamic-oxaloacetic transaminase | 10% | 10% | 10% | 20% | 20% | 20% | 100% |
| ultrasonography | 20% | 20% | 20% | 20% | 20% | 20% | 100% |
| contact tracing visit | 0% | 0% | 0% | 0% | 0% | 0% | 0% |
| magnetic resonance imaging | 5% | 0% | 10% | 10% | 10% | 10% | 10% |
| tracing those who are lost to follow-up | 20% | 20% | 20% | 20% | 20% | 20% | 20% |
| patient support costs | 0% | 0% | 0% | 0% | 0% | 0% | 0% |
| active TB drug safety monitoring and management | 100% | 100% | 100% | 100% | 100% | 100% | 100% |
| adverse event management through IP care | 5% | 5% | 5% | 5% | 5% | 5% | 10% |
| elective partial lung resection (lobectomy or wedge resection) | 1% | 0% | 0% | 5% | 5% | 5% | 50% |
| follow-up post-TB treatment | 90% | 90% | 90% | 90% | 90% | 90% | 90% |
| Post TB lung disease care | 10% | 0% | 10% | 20% | 20% | 20% | 50% |
| Palliative care | 5% | 50% | 5% | 10% | 50% | 50% | 50% |

#### Quantity and Population in Need (PIN) of prevention interventions/service

*Table 15: TB preventive treatment related interventions/services (quantities)*

| **Intervention /service** | **DS: 3 HR (pediatric)** | **DS:3 HP (adult)** | **DR: 6 levofloxacin daily (pediatric)** | **DR: 6 levofloxacin daily (adult)** |
| --- | --- | --- | --- | --- |
| patient support costs | 1 | 1 | 1 | 1 |
| directly observed treatment | 1 | 1 | 1 | 1 |
| liver function test | 1 | 1 | 1 | 1 |
| serum glutamic pyruvic transaminase | 1 | 1 | 1 | 1 |
| patient counselling | 1 | 1 | 1 | 1 |
| In patient management of Drug sensitive TB | 1 | 1 | 1 | 1 |
| contact tracing visit | 1 | 1 | 1 | 1 |
| patient support costs | 1 | 1 | 1 | 1 |

*Table 16: TPT related interventions/services (PINs)*

| **Intervention abbreviation** | **DS: 3 HR (pediatric)** | **DS:3 HP (adult)** | **DR: 6 levofloxacin daily (pediatric)** | **DR: 6 levofloxacin daily (adult)** |
| --- | --- | --- | --- | --- |
| patient support costs | 0% | 0% | 0% | 0% |
| directly observed treatment | 10% | 10% | 10% | 10% |
| liver function test | 5% | 5% | 5% | 5% |
| serum glutamic pyruvic transaminase | 5% | 5% | 5% | 5% |
| patient counselling | 1% | 1% | 1% | 1% |
| In patient management of Drug sensitive TB | 1% | 1% | 1% | 1% |
| contact tracing visit | 0% | 0% | 0% | 0% |

### **References**

| [1] | A. Health., "https://avenirhealth.org/software-spectrum.php. 2023," 2023. [Online]. |
| --- | --- |
| [2] | J. Fox, "Contact investigation for tuberculosis: a systematic review and meta-analysis," *Eur Respir J,* vol. 41, no. 1, pp. 140-56, 2013. |
